# Epidemiological monitoring and control perspectives: application of a parsimonious modelling framework to the COVID-19 dynamics in France

**DOI:** 10.1101/2020.05.22.20110593

**Authors:** Mircea T. Sofonea, Bastien Reyné, Baptiste Elie, Ramsès Djidjou-Demasse, Christian Selinger, Yannis Michalakis, Samuel Alizon

## Abstract

SARS-Cov-2 virus has spread over the world creating one of the fastest pandemics ever. The absence of immunity, asymptomatic transmission, and the relatively high level of virulence of the COVID-19 infection it causes led to a massive flow of patients in intensive care units (ICU). This unprecedented situation calls for rapid and accurate mathematical models to best inform public health policies. We develop an original parsimonious model that accounts for the effect of the age of infection on the natural history of the disease. Analysing the ongoing COVID-19 in France, we estimate the value of the key epidemiological parameters, such as the basic reproduction number 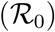, and the efficiency of the national control strategy. We then use our deterministic model to explore several scenarios posterior to lock-down lifting and compare the efficiency of non pharmaceutical interventions (NPI) described in the literature.

## 1 Introduction

### 1.1 The COVID-19 pandemic

In Dec 2019, a rapidly increasing number of ‘pneumonia of unknown etiology’ cases in Wuhan, China, triggered a specific surveillance mechanism implemented by China’s Center for Disease Control (CDC) in the wake of the 2003 SARS-CoV pandemic (Li et al., 2020a). In Jan 2020, China CDC identified the new inter-human transmitted pathogen as a coronavirus, which was later named SARS-CoV-2 after its phylogenetically close predecessor (Coronaviridae Study Group of the ICTV, 2020). Meanwhile, the respiratory disease for which it is responsible, called COVID-19, was better described from the clinical point of view, especially by identifying age and co-morbidities as risk factors of symptomatic aggravation and fatality (Chen et al., 2020; Huang et al., 2020). Because of its relatively high basic reproduction number 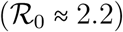 in the Wuhan outbreak (Li et al., 2020a) and the high transimissibility of asymptomatic/undocumented cases (Nishiura et al., 2020a; Li et al., 2020b), as well as the intensity of international travel, the virus rapidly spread in mainland China and then all over the world.

On Mar 11, 2020, the WHO announced that the COVID-19 outbreak had reached pandemic stage. To date, no pharmaceutical treatment has proven to be efficient, neither prophylactically nor therapeutically.

### 1.2 The French situation

The documented importation of SARS-CoV-2 on the French metropolitan territory dates back to Jan 24, 2020 with the detection of three cases with a travel history to Wuhan (Stoecklin et al., 2020). These were also the first COVID-19 cases detected in Europe.

From Feb 27, the daily incidence of detected cases started to increase exponentially, followed on Mar 8 by the daily hospital mortality. On Feb 29, the government announced that France had moved to stage 2 of the epidemics with isolated cases, and reminded that masks should only be worn based on medical advice. On Mar 7, the government advocated for basic measures (hand washing, avoiding handshakes) but announced the elections on Mar 15 were not cancelled. On Mar 12, a scientific council was set up and the president announced that from Mar 16, all schools and universities would be closed, before implementing a lock-down for the whole country from Mar 17. 55 days later, on May 11, the lock-down was lifted.

### 1.3 COVID-19 epidemic modelling

Mathematical modelling was involved early on to estimate the magnitude of the COVID-19 epidemic in Wuhan. Estimates of the serial interval were obtained rapidly allowing to estimate the basic reproduction number (Li et al., 2020a). Later on, analyses on the number of travelers from Wuhan to Europe and infected by COVID-19 allowed to estimate the magnitude of the epidemics in Wuhan (Imai et al., 2020) or the incubation time of the infection (Backer et al., 2020). Furthermore, stochastic models allowed to better assess the probability to control initial outbreaks (Hellewell et al., 2020). Early models also stressed the importance of detecting infections early on to control the epidemics (Ferretti et al., 2020).

It quickly became clear that the epidemic had reached a large enough size to escape stochastic forces in Wuhan and that, in spite of a full lock down and travel restrictions, it had spread all over the world, making deterministic models more appropriate. This also led to an increase in modelling efforts because deterministic compartmental models based on ordinary differential equations are more commonly used in epidemiological modelling. However, although these models have useful analytical properties when analysed over a long time period, they are perform poorly on short time scales. One reason for this is that they are essentially Markovian or ‘memoryless’. This means for instance that an individual that has been infected for 10 days has the same probability to clear the infection that someone infected for less than a day. Over a long time period this effect averages out but on a shorter time scale, even if the dynamics are deterministic, this Markovian assumption makes it difficult to reconcile the model with data.

Focusing on the French epidemic, Di Domenico et al. (2020) developed a variant of an SEIR model with multiple levels of infection severity and estimated the 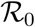 to 3.0 (with a 95% CI of 2.8-3.2). Using an SIR model, Magal and Webb (2020) estimated it to 4.5 (no confidence interval computed). Using a meta-population model, Roux et al. (2020) estimated it in various regions of France and found values between 1.7 and 4.2 with a median value of 2.8. Using a simpler SIR model but a more robust statistical framework Roques et al. (2020), found an 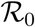 between 3.1 and 3.3, but also estimated a temporal reproduction number 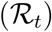 after the lock-down of 0.47 (with a 95% CI of 0.45-0.50). Hoertel et al. (2020) developed a detailed agent-based simulation model with 194 parameters, which allows them to estimate 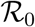 to 3.1 (no confidence interval computed) and investigate the effect of different types of interventions on the spread of the epidemics. Finally, Salje et al. (2020), with priority access to restricted ICU patient data, developed a detailed SEIR model with explicit ICU admissions tailored to the French epidemics. Using a detailed statistical inference model, they estimated the 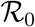 to 2.9 (with a 95% CI of 2.8-2.99) and an 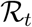 after the lock-down of 0.67 (with a 95% CI of 0.65-0.68).

The model we introduce is inspired by an earlier optimal control model (Djidjou-Demasse et al., 2020). Taking advantage from its two key qualities, namely being simultaneously deterministic and tailored for non-exponentially distributed transition times, accurately follow the main properties of COVID-19 epidemic, while limiting the number of compartments and parameters. In this study, by using the French COVID-19 epidemics as an example, we introduce a more mechanistic compartmental model, which explicitly features two categories of critically ill patients. This allows us not only to describe the past state of the epidemic but also to explore the effect of original and ea control strategies.

### 1.4 A non-markovian discrete-time model

Unlike the majority of deterministic models used in epidemiology (Keeling and Rohani, 2008), ours runs in discrete time, with a time step equal to one day. This choice implies a formalism of numerical sequences that is closer to algorithmic syntax, less compact and less intuitive than that of ordinary differential equations. However, discrete-time models have the great advantage of implementing process memory. Indeed, the usual continuous-time models are said to be memory-less because the probability for an individual to leave a certain compartment, for instance to recover, is independent of the time already spent in this compartment.

The absence of memory does not alter the qualitative properties of the model when an asymptotic behaviour is studied. It can however become extremely oversimplifying when studying phenomena over short time scales, that is in the transient regime of the system. Indeed, failing to account for the past may buffer sudden changes in the system. In the case of COVID-19 infections, it is very rare for symptoms to appear the day after contamination. On the contrary, clinical signs appear mostly at the turn of the week following the contamination. Failure to take this into account affects the short-term dynamics and, as we show, make it impossible to interpret the incidence data.

Although there are ways to correct ordinary differential equation models to deal with each of the above-mentioned situations (e.g. delays, impulses, non-autonomous equations), the discrete-time approach makes it possible to easily accumulate all of these options. It also approaches the very format of the data collected, monitored, and communicated during epidemics on a daily basis.

While accounting for non-Markovian transition times between clinical-epidemiological compartments, our model remains deterministic. This means that contrarily to agent-based models, it can explore a variety of scenarios in a computationally efficient way. It thus combines the advantages from both stochastic modelling (namely simulating non-exponential-like waiting times) and law of large numbers (namely capturing the general trend of the system by averaging). Moreover, the deterministic nature of the model makes it quite parsimonious, allowing unknown parameter values to be estimated, thereby preventing arbitrary choices. Likewise, its simple formulation opens ways to a variety of extensions and facilitates broad applications to health authorities.

### 1.5 Peak dynamics and epidemic control

By calibrating our model using French data, we show that epidemiological dynamics can be accurately captured. More precisely, we estimate the 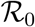 of the French epidemics to be between 2.59 and 3.39, a number that decreased to between 21.3 and 27.1% of its value after the lock-down (95% likelihood interval).

By comparing our calibrated model to one without memory, we show the importance of allowing model parameters to depend on the age of the infection to accurately capture short-term dynamics. We also investigate the effect of earlier or later implementation of a national lock-down on epidemic peak intensity and timing.

Finally, we use our flexible and statistically robust framework to compare 3 main types of non-pharmaceutical interventions (NPI) that have been proposed in the literature: focusing the control on sub-populations who are more at risk, implementing periodic control, and deploying an adaptive lock-down (also known as ‘stop and go’). We show that these differ in terms of cumulative mortality but also in terms of the cumulative number of people undergoing strict control. This opens new perspectives for NPI, which are essential until pharmaceutical options become available.

## 2 Methods

### 2.1 Model structure and dynamics

Following classical epidemiological models (Kermack and McKendrick, 1927; Keeling and Rohani, 2008), we group individuals with the same contribution to the dynamics into compartments whose densities are tracked over time. We consider age structured dynamics, by splitting each compartment into an arbitrary number of age groups, which are hereafter denoted by an index *i*. This enhances the model with two key features. First, many COVID-19 clinical parameters are age-dependent, especially the infection fatality rate (Verity et al., 2020a). With this age structure, we can adjust nationwide averages and capture demographic effects by matching demographic data to age-stratified medical data. Second, we can investigate age-differentiated non-pharmaceutical interventions (NPI). Note that this model can be extended to formally take into account any kind of finer stratification, e.g. age, sex and comorbidities simultaneously. Furtermore, adding a discrete implicit spatial (also known as meta-population) structure is straightforward.

Since hospital admissions and critical cases are less frequent than non-severe infections, we assume that all transmission events occur in the community. A specific extension of the model could be to focus on nosocomial transmission.

The structure of the system is shown in Figure 1. Initially, all individuals in group *i* belong to the susceptible compartment, the density of which is denoted *S_i_*. These individuals can be infected with a probability Λ*_i_* called the force of infection (the *i* subscript indicates that not all ages are equally susceptible to infection).

**Figure 1:**
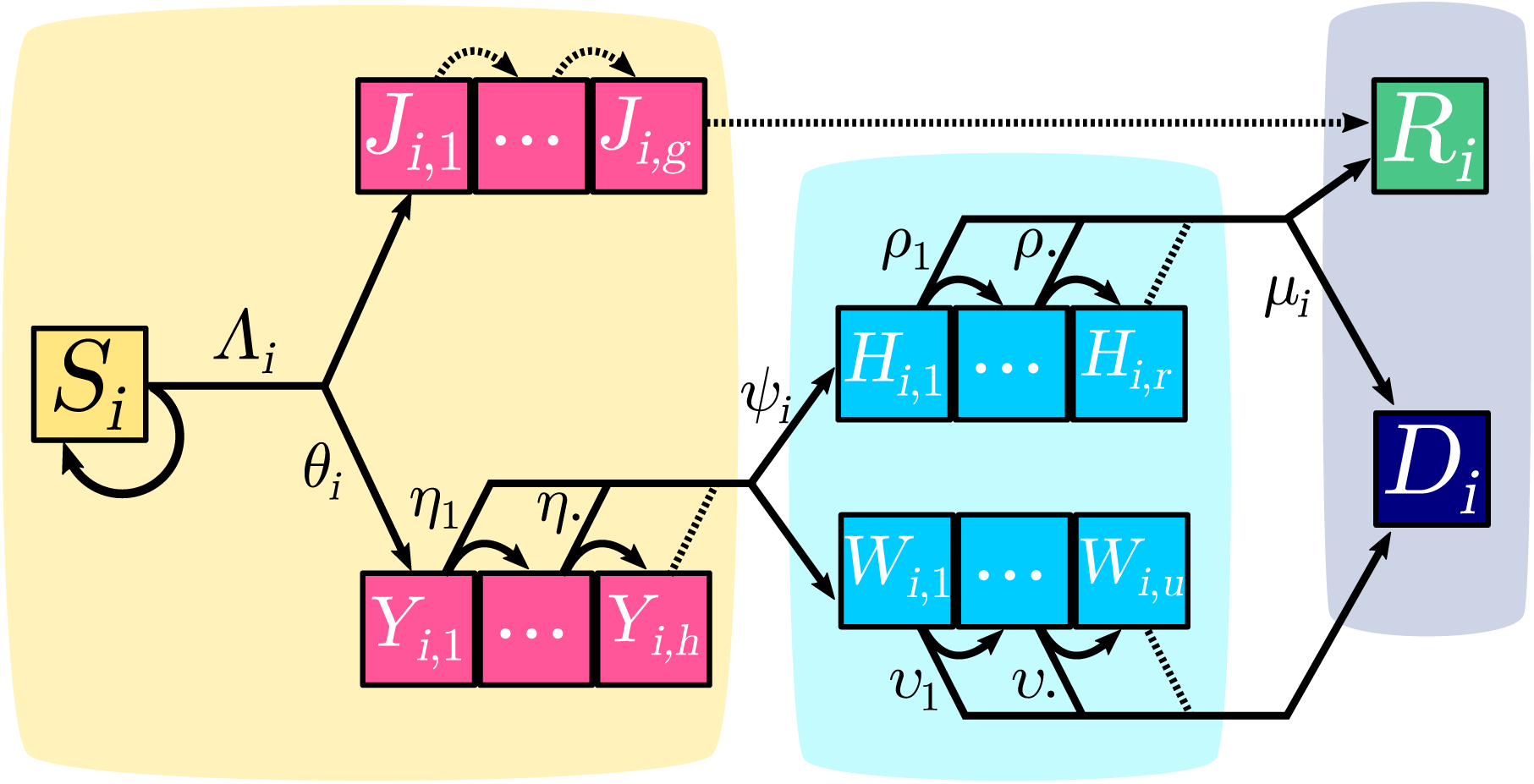
COVID-19 epidemic discrete time model structure. Each square represents a group of individuals who share the same clinical kinetics and who contribute equally to the epidemic dynamics. Contiguous squares form a compartment, in which each individual progresses day after day, therefore allowing to capture memory effects of the infection age. Pink boxes correspond to infected individuals in the community (depicted by the yellow area). Light blue boxes represent the hospitalized critical cases (the light blue area depicting the hospital). The purple-grey area corresponds to removed compartments that do not contribute to the epidemic. Arrows between boxes correspond to the daily flow of individuals from one compartment to the other. Dotted arrows depict transitions that occur with probability 1. The *i* subscript indicates the age group. For the sake of simplicity, only one group is depicted here and only one of the two probabilities is shown for each bifurcating transition (the other being its complementary to 1). Details about the compartments, flows and notations are provided in the main text.

Upon infection, a fraction 1 − *θ_i_* of the individuals will develop non critical infections and move to the *J_i,_*. compartment, where the second subscript indicates the age of the infection (in days). At each time step, an individual in the compartment *J_i,k_* moves to the compartment *J_i,k_*_+1_ and after *g* days of infection it moves to the recovered (and assumed lastingly immunized) compartment *R_i_*.

A fraction *θ_i_* of infections will lead to critical complications (typically acute respiratory distress syndrome (Bouadma et al., 2020)), and move to the *Y_i_*_,1_ compartment. Every day *k*, individuals in the *Y_i,k_* subgroup have a probability *η_k_* to be hospitalized and a complementary probability 1 − *η_k_* to move to the *Y_i,k_*_+1_ subgroup. No individual of a cohort of critical cases remains in the *Y_i,_*. compartment after *h* days.

As detailed in Appendix S2.5, two groups of hospitalized critical patients are considered. Those who have a substantial chance of recovering and who will benefit from a long stay (at least greater than one day) in an intensive care unit (ICU), are denoted by *H_i,_*.. Those who will die, either after a short stay in ICU or in another ward are denoted by *W_i,_*.. Upon hospitalization, a proportion of the incoming *Y_i,_*. moves to *H_i,_*. while the remaining part moves to *W_i,_*.. Time to death for the latter compartment is captured by the sequence of *υ_k_*. For the former, ICU discharge occurs with probability *ρ_k_*, *k* being the number of days after hospitalization, and only a fraction 1 − *μ_i_* of individuals survive.

### 2.2 Forces of infection

In the SIR-like modelling framework, the force of infection refers to the infection rate per capita of susceptibles, often expressed as *λ*: = *βI* (Keeling and Rohani, 2008). Equivalently, the instantaneous incidence is *βIS = λS*, which is the translation of the mass action law implied by the mean-field approximation made by such spatially unstructured models.

In our discrete time model, the force of infection Λ*_i_* is not a rate but a daily probability of infection (*per capita* of susceptibles from group *i*) that saturates with the prevalence. Individual contributions of infected individuals are not additive when prevalence is high because a susceptible host surrounded by infected individuals can be infected by several of them the same day. When prevalence is low, the probability of contamination by multiple infectors the same a day is low and the force of infection is well approximated by the sum of contributions of each infected individual. Λ*_i_* is therefore a monotonically increasing function of prevalence, bounded by 1 and with a positive initial slope.

As we show in Appendix S2.2, deriving an expression for the force of infection is far from trivial for two reasons. First, we do not have a single class of infected individuals *I* as in most simple models. We therefore need to define and calculate an effective infectious density at time *t* (denoted 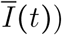, which can be seen as the number of individuals in the *J* and *Y* classes, weighted by both the level of per-capita contact ratio *c* (*t*) and the generation time distribution, here approximated by the serial interval.

As detailed in Appendix S2.2, we find that the force of infection can be written as

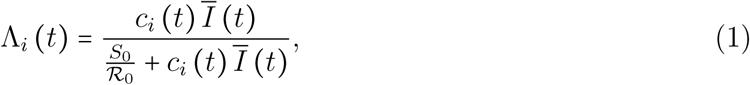

where *c_i_* denotes the relative contact rate per capita of *i*-individuals (i.e. the current contact rate divided by the pre-epidemic baseline), 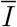 the effective infectious density, *S*_0_ the initial population size and 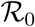 the basic reproduction number. In Appendix S2.2, we also demonstrate the origin of the saturation parameter in the Michaelis-Menten equation, namely 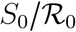.

### 2.3 Data

This model can be applied to any population where COVID-19 incidence data can be collected. However, it also requires additional details, such as the time spent in ICU or the mortality, which is why we apply it to France where at least part of the data is available.

All the model parameters are detailed in Appendix S1. We attempted to calibrate the models to the best of our ability based on the data available at the time of writing this report. For instance, the calculation of *μ_i_* is based on reports from Santé Publique France. The age structure of the population is based on the data from the French National Institute of Statistics and Economic Studies (Institut National de la Statistique et des Études Économiques, 2020).

Due to the lack of accessible public health data from France, we had to combine it with data from other countries. We therefore approximated the generation time using the serial interval inferred by Nishiura et al. (2020a) on transmission pairs from multiple countries, as well as the Infection Fatality Ratio (IFR) computed by Verity et al. (2020a).

Further details about the time series data can be found in Appendix S2.4.

### 2.4 Memory effects

For several key processes in the model, the probability for an event to occur depends on the elapsed time. This is the case for the interval between contamination and hospitalization, through the probability distributions that underlie the *η_k_* sets of parameters. This is motivated by the fact that, as early noticed by clinicians (Bouadma et al., 2020), respiratory complications of COVID-19 arise in a quite narrow time window approximately a week after symptom onset.

Memory is also involved in the transmission term. As one can notice, there is no incubation period *per se* in the model. This is implicitly accounted for in the infectiousness term *ζ_k_* used to calculate the force of infection (see Appendix 2.2 for details). Indeed, the compartments that transmit the virus (*J* and *Y*) do so following weights, which we refer to as the discretized generation time, i.e. the daily probability of transmitting the virus each day following the day of infection. Based on empirical data, e.g. the serial interval derived by Nishiura et al. (2020a), the first day these values are very low, then they increase rapidly before decreasing slowly. This modelling approach eventually recovers a classical SIR model for the non-critical cases, capturing the empirical and effective non-homogeneous infectiosity period without recourse to additional compartments (such as a latent/exposed *E*, and convalescent densities) and their corresponding parameters.

We use two parameters to describe non-exponential distribution, which we assume to be Weibull distributions with a shape parameter greater than 1 (which captures the ‘ageing’ property) and a scale parameter. Other components of the model could also, in theory, accommodate memory processes, namely the time distribution from hospitalization to ICU discharge or death (*ρ_k_* and *υ_k_*). However, preliminary fitting attempts reveal that an exponential (memory-less) distribution is more parsimonious. In Appendix S2.6, we explain in further details how the distributions for the various waiting times were defined and estimated from the data.

For comparison purposes, we also considered the continuous-time memory-less translation of this model (detailed in Appendix S2.9), which proved to be less adequate to capture the dynamics of the French epidemic in France, as shown by Fig. 3.

### 2.5 Parameter estimation and simulation confidence intervals

Parameter estimation is required prior to analyzing the outcome of the model when i) strong simplifying assumptions about the phenomenon under study are made, and ii) some parameter values are not known with sufficient certainty.

First, the strongest assumption of the present model is the mean-field approximation: the population is supposed to be well-mixed, which is obviously not the case at the country scale. Nonetheless, earlier works have shown that non spatially structured models produce conservative estimates from a public health viewpoint, while their parsimony and tractability outweighs the greater precision provided by finer models (Keeling, 1999; Trapman et al., 2016).

Second, the value of the basic reproduction number, of the day of onset of the epidemic wave, or of the lock-down effect have only been estimated in recent modelling works (ETE Modelling Team, 2020b; Danesh et al., 2020; Salje et al., 2020). Those values might not directly apply to our model and corrections might be needed. Likewise, known COVID-19 generation time distributions (Nishiura et al., 2020b) or IFR (Verity et al., 2020b) originate from field studies outside France. Besides, the estimated distribution of the ICU admission to death interval does not capture reporting delays (note that deaths and ICU admission are not coming up through the same channels). Parameter fitting is thus used to account for the uncertainty in initial parameter values, thereby improving predictions by re-calibrating their value.

Parameter inference was performed using nationwide daily ICU admissions, current ICU bed occupancy, as well as the cumulative number of deaths, all published by Santé Publique France and available on the French government data repository (Santé Publique France, 2020b).

We first located the region of highest likelihood using initial parameter values estimated from data or compatible with the literature. Then, the maximum likelihood estimates (MLE) and associated 95%-intervals were calculated stepwise with respect to daily ICU admissions, ICU discharges, and finally daily mortality time series.

Confidence intervals for the simulation outputs are based on a collection of parameter sets assumed to be equally likely. These parameter sets originate from random draws according to a multivariate Gaussian distribution (centered around the maximum likelihood parameter set and variances based on the confidence interval of each parameter) and only the resulting parameter sets whose likelihoods are not significantly different from that of the MLE are kept. The 95% confidence intervals of the model’s output are then calculated as the daily sample quantiles across all runs.

Further details about the procedure used for parameter estimation can be found in Appendix S2.7. The methods used to obtain predictions of the dynamics are shown in Appendix S2.8

## 3 Results

### 3.1 Epidemic parameter values estimation

By analysing nationwide hospital data, publicly provided by Santé Publique France (Santé Publique France, 2020b), we obtain maximum likelihood estimates for all parameters except those determining the generation time distribution, that was kept fixed following Nishiura et al. (2020a).

The estimates and their likelihood intervals are summarised in Table S-5. The basic reproduction number is 2.99 (95% likelihood interval: [2.59-3.39]), consistent with most estimates for the French epidemics. The effect of the lock-down is estimated to a 75.9% (72.9-78.7) reduction of the reproduction number. The estimates for the other parameters are all in line with official reports of average values (we cannot have access the exact distributions yet). The epidemic wave is estimated to have originated around Jan 20, in agreement with early phylogenetic analyses on sequence data (Danesh et al., 2020).

The fitted non-markovian discrete time model accurately captures the dynamics of both the daily hospital mortality and the daily number of ICU admissions since most of the data points fall into the 95% confidence intervals. As can be observed in Figure 2(top), the model correctly approaches the number of daily admissions in the vicinity of the peak, which is crucial for hospital management at the local but also national level.

**Figure 2:**
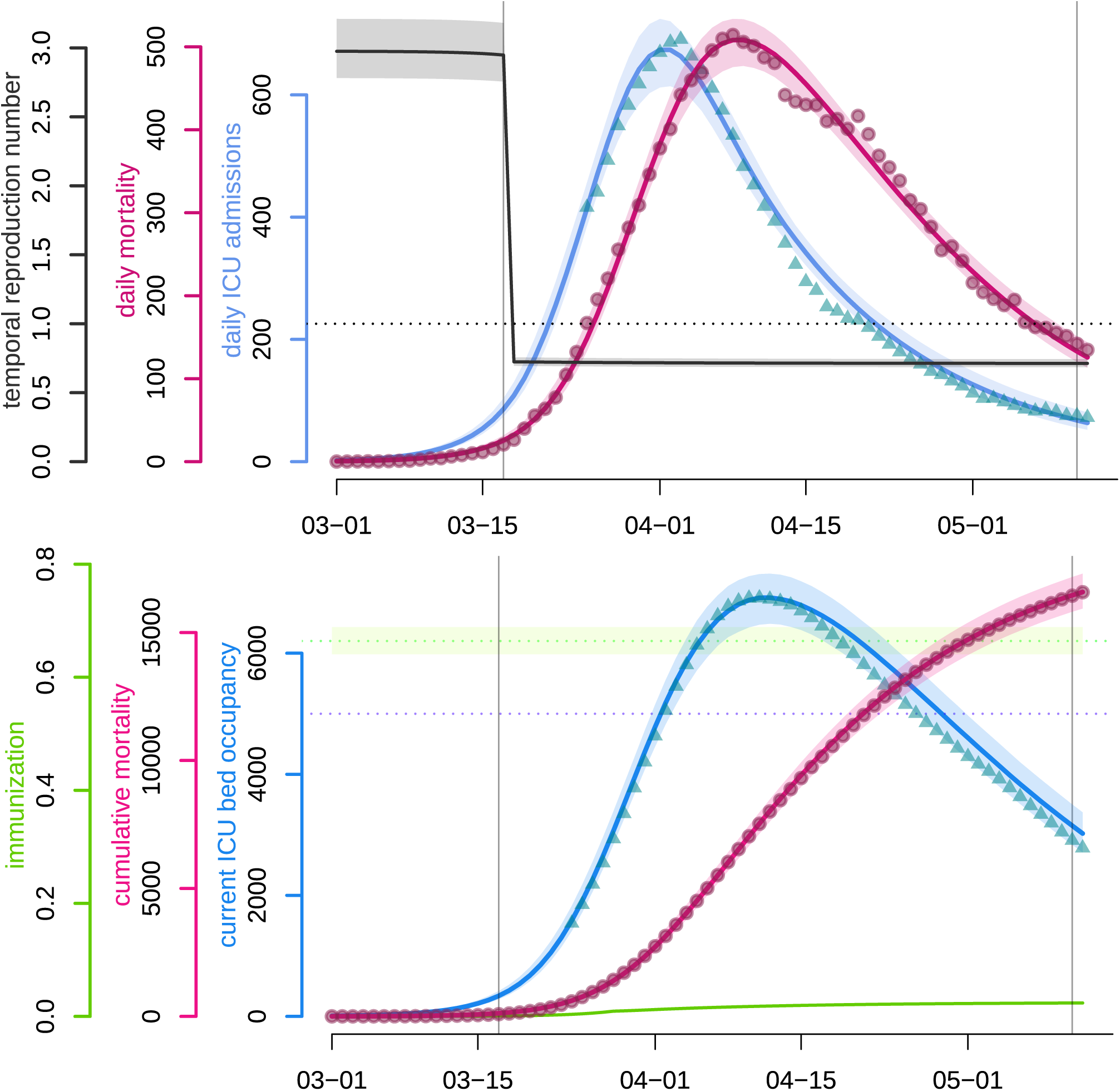
COVID-19 epidemic wave in France as fitted by a non-markovian discrete time model. **Top panel**. The blue and pink curves respectively represent the median daily ICU admissions and the median daily (hospital) mortality as generated by the fitted model.Turquoise triangles and red circles are the (rolling 7-day average) data counterparts. The black curve shows the median daily temporal reproduction number calculated from the simulated epidemic. The dotted horizontal line shows the reproduction number threshold value, i.e. 1. **Bottom panel**. The blue and pink curves respectively represent the median number of occupied beds in ICU nationwide and the median cumulative (hospital) mortality as generated by the fitted model. The turquoise triangles and red circles are the (rolling 7-day average) data counterparts. The purple dotted horizontal line shows the initial French ICU capacity, ca. 5,000 beds. The green curve shows the median proportion of the population that has recovered (and is assumed to be immune). The green dotted horizontal line corresponds to the median herd immunity threshold. The two vertical lines show respectively (from left to right) the beginning and the end of the French national lock-down. Shaded areas correspond to 95% confidence intervals.

We also present the temporal reproduction number 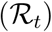, which rapidly drops below unity following the onset of the national lock-down and was equal to 0.71 [0.69,0.74] by May 11 according to the model. Notice that 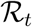 started to decrease before the lock-down onset, due to density dependent effects. In a model with strong host heterogeneity and so-called ‘super-spreaders’, this effect would be even more pronounced. Other explanations include local saturation, staggered implementation of pre-containment measures, such as health communication campaigns, or improved patient management as diagnosis and therapy became more effective. However, neither the structure of the model nor the level of detail in the available data makes it possible to identify the isolated impact of each measure on epidemiological dynamics.

Figure 2(bottom) illustrates that the model also accurately captures the post-ICU admission dynamics (though with a slight tendency to overestimate the declining ICU bed occupancy), which is essential in assessing the risk of a saturation of such hospital units, which would lead to an excess-mortality. The fitting of these data points could be improved with access to non-aggregated patient data or distributions of ICU residency time. The cumulative mortality curve is fitted with great accuracy, which allows us to use it as a comparison criteria between control strategies in further analyses. Finally, the figure also shows that the level of population immunisation, the median of which we estimate at 2.37% ([2.27,2.48] % 95%-CI) by May 11, which is far below the classical group immunity threshold.

We also performed the parameter value inference by censoring the data to the right in order to assess the relevance of estimates obtained earlier in the epidemics. As shown in Appendix S3.2, estimates with a censoring on Apr 15, that is one month before the final data point shown in Figure 2, were already accurate. Estimates with an earlier censoring are qualitatively correct but the confidence intervals larger.

To illustrate the ability of our discrete model to capture COVID-19 short-term dynamics, we show in Figure 3 the best outcome of parameter inference for a Markovian (memory-less) model. In spite of one additional degree of freedom compared to the focal model (see Appendix S2.9 for more details), the best fitted curves with the memory-less model (respectively in blue and pink) fail to capture the timing and amplitude of the peaks, while their decline is slower than the data.

**Figure 3:**
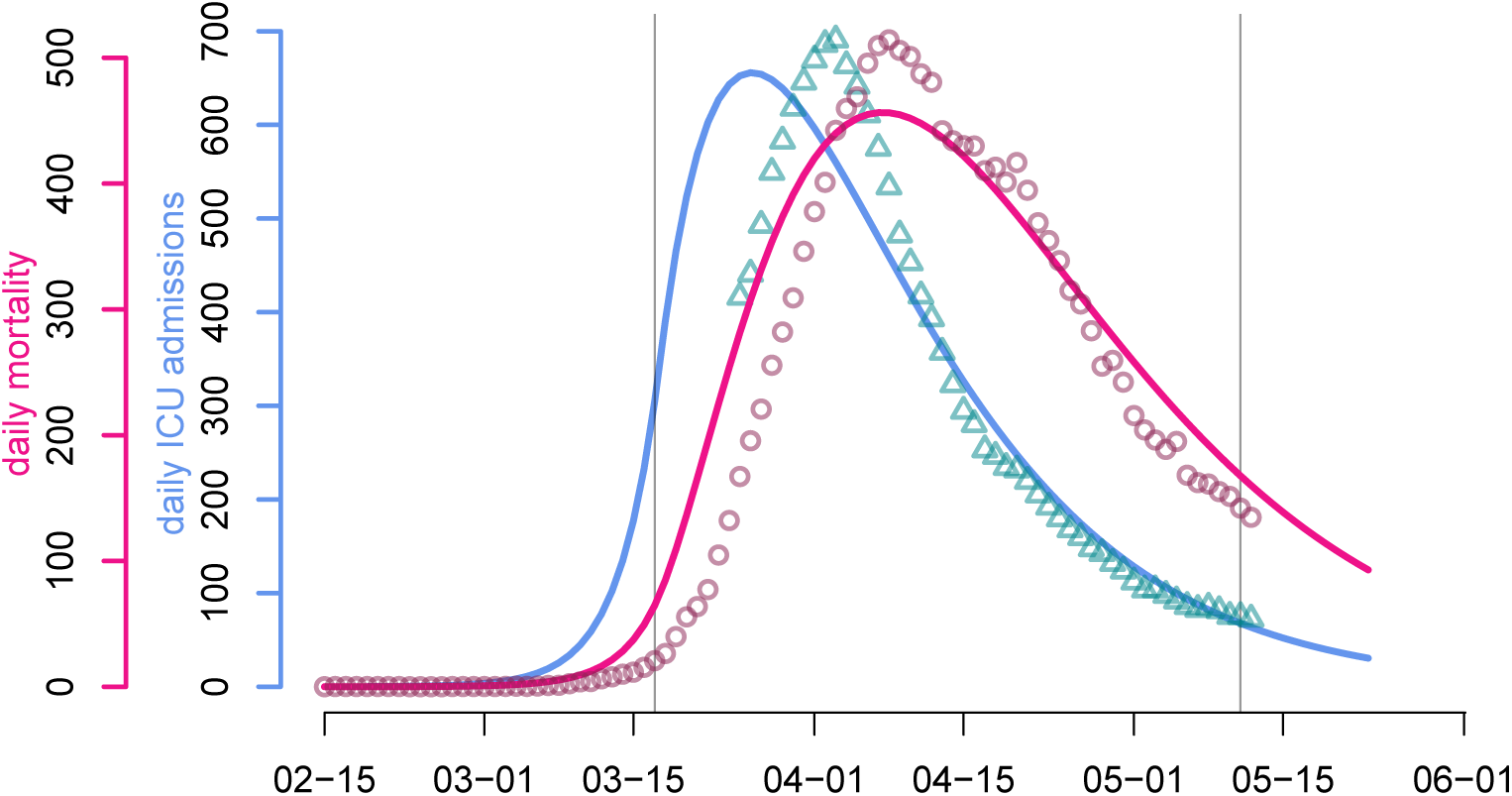
Predicted (plain line) and observed (dots) dynamics with (classical) memory-less processes. The continuous-time memory-less analog of the focal model poorly reproduces the observed trends in daily ICU admissions (turquoise triangles) and daily mortality (red circles).

### 3.2 Response date impact

Using our estimated parameters, we then explored the effects of implementing the national lock-down a week earlier or a week later. As shown in Figure 4, the peak was reached on Apr 8, with 7019 ICU beds occupied (the model estimates it to Apr 12 and 6920 beds). Enforcing the lock-down a week earlier (in green) would have led to an earlier and smaller epidemic peak with less than 1,500 ICU beds occupied on Apr 5. Conversely, another week of delay (in red) would have led to a peak above 32,000 beds occupied in ICU on Apr 18, which is largely above the ICU capacity at the time (approximately 5,000 beds). Overall, in the range studied, each elapsed week multiplies ICU occupancy peak by more than 4.5, while delaying it by 3 weeks.

**Figure 4:**
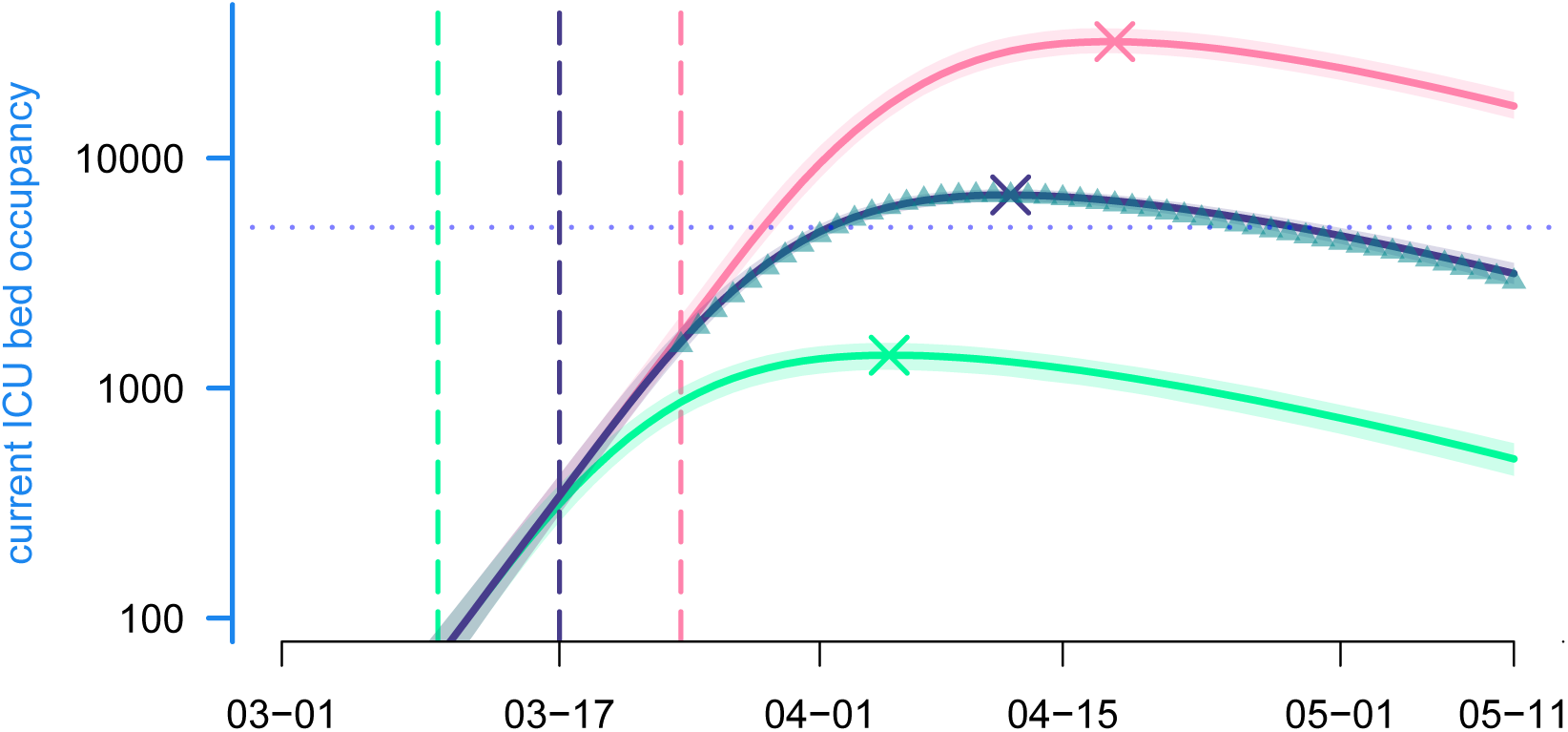
Lock-down implementation date effect and ICU bed occupancy. Each curve represents the median current ICU bed occupancy as generated by the model according to a given lock-down scenario, while their surrounding shaded areas correspond to the their 95% confidence interval. From bottom to top: the green scenario simulates an early national lock-down (on Mar 10th); the purple scenario is the realised one (lock-down beginning on Mar 17); the pink scenario simulates a late lock-down (on Mar 24th). Vertical lines indicate lock-down implementation dates. Crosses indicates the median ICU peak activity. Triangles represent the data and the dotted line the maximum ICU bed capacity in France.

These differences also translate in terms of mortality. Implementing the lock-down a week earlier could have led, according to the model, to 13,300 [12,900-13,700] less deaths, while waiting for an additional week could have claimed 52,800 [45,800-61,500] more lives (Figure S-6).

### 3.3 Predicting future dynamics

This model cannot estimate the effects of the end of the national lock-down in France on May 11, as it relies on data notifying events that occur on average two (ICU admission) to four (death) weeks after contamination. However, assuming our estimated value hold after the lock-down is ended, we can explore the future dynamics as a function of the post-lock-down reproduction number. The aim is not to make predictions, since there is no data to test them, but rather to highlight the interplay between the post-lock-down reproduction number and NPI-enforcement timing with respect to a potential epidemic rebound. For illustration purposes, the chronicles of four scenarios are provided in Appendix S3.4.

We also provide a graphical online interface (provided as a supplementary data) to maximise the reach of the model.

Following a more systematic approach of end of the national lock-down, we investigated the effect of the NPI reinforcement delay on the ICU peak height and timing. The results are shown in Figure 5. As expected, a reproduction number less or equal to 1 (in green) does not require any control reinforcement. Conversely, higher values of 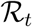 trigger an epidemic rebound, that can saturate France’s ICU capacity (ca. 5,000 regular beds) with values as low as 1.2 [1.1, 1.3] if an appropriate control response is not implemented before mid-July. For 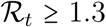, we find that a reinforcement as early as mid-June is necessary to preserve the national health system (Figure 5, top panel). Additionally, Figure 5 (bottom panel) shows that if a massive peak is to occur 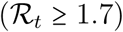, it will likely be in the second part of July, even if an appropriate response in mounted in mid-June or later. For lower peaks 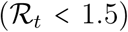, the height of which can be substantially reduced by timely reinforcement of control, occur within one month (early responses) to two weeks (late responses) after NPI implementation.

**Figure 5:**
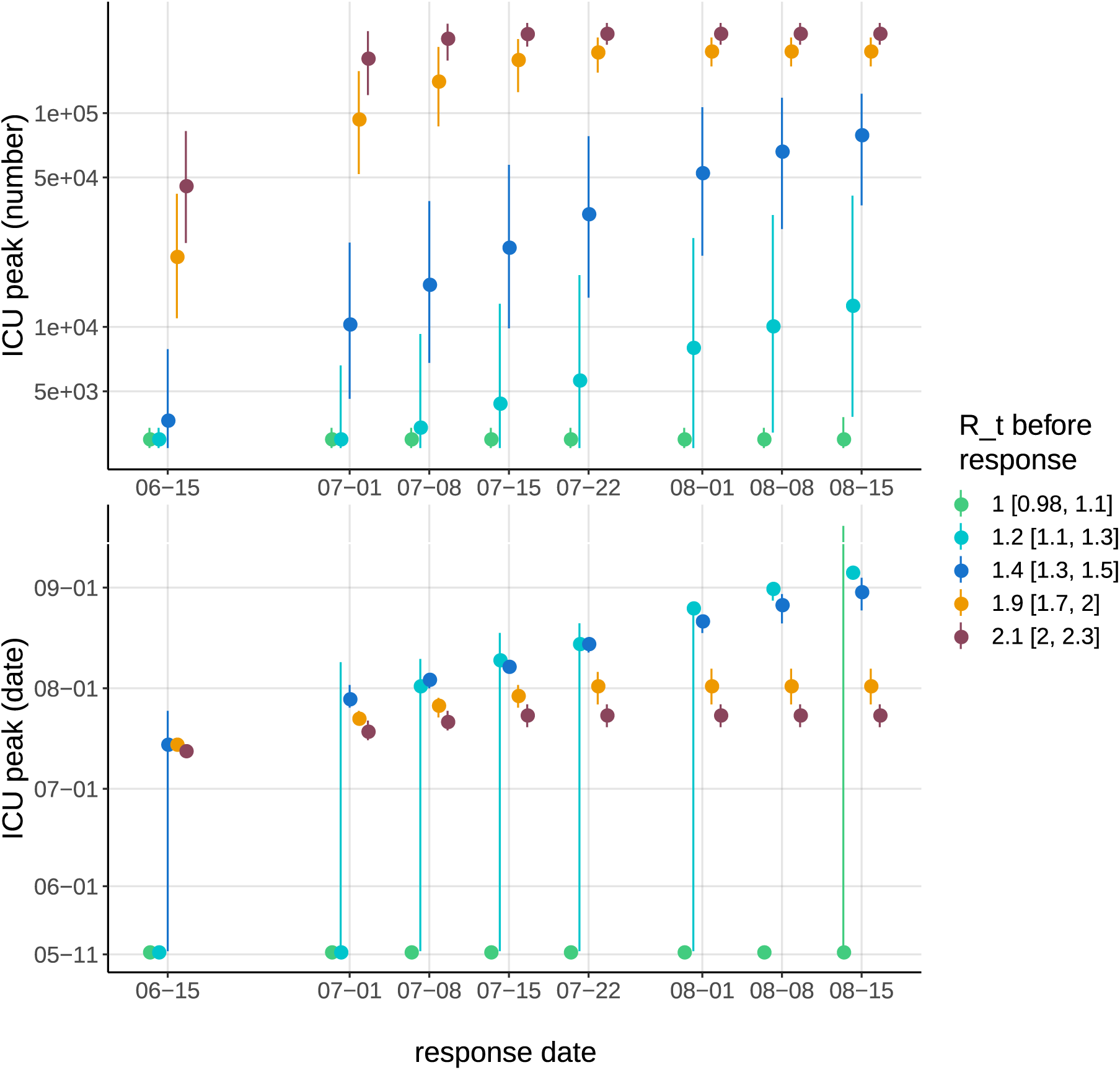
Effect of reproduction number and NPI response on ICU peak dynamics. Colors indicate post-lock-down reproduction number, which ranges from 1 (green) to 2.1 (brown) (see the legend with the 95%-CI). The abscissa indicate the date of implementation of renewed NPI that bring the reproduction number to 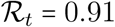 [0.85,0.98]. Each dot represents the median highest number of ICU occupied beds (top panel) and the median date at which the peak is reached (bottom) (the bars indicate the 95% CI). Dots at the bottom of each panel correspond to an absence of epidemic rebound (therefore the peak is artificially considered to be on May 12). The vicinity with the threshold value 1 for 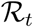 explains the large CI of the peak dates for the green and turquoise scenarios.

### 3.4 Comparing intervention strategies

In this section we use the model previously fitted on the data available as of May 12 to explore control strategies that belong to three main classes of non-pharmaceutical interventions (NPI): adaptive (indicator-triggered) lock-down, periodic alternation of lock-down and release phases (not indicator-triggered), and age-differential control.

We compare scenarios using the predicted cumulative hospital mortality on Dec 31 2020. The choice of this criterion is motivated by three reasons. First, the mortality time series is the one captured with the greatest accuracy by the model. Second, in the absence of pharmaceutical solutions, deaths are a more stable indicator of the state of the epidemic (e.g. the timing of hospitalizations may change as knowledge of the disease accumulates). Third, peak ICU activity and concern is more likely to vary among countries, which makes it less generic.

Importantly, the following simulations are in no way intended to be statistical forecasts of the COVID-19-related death toll at the end of 2020 in France or elsewhere, but rather a numerical illustration of the non-trivial interplay between the degrees of freedom in each of the NPI strategies considered. For the sake of parsimony, we set the implementation of all further considered strategies by May 12, thus avoiding to consider multiple scenarios between lock-down lifting and new NPI reinforcement (this aspect having been addressed above).

To facilitate the comparison in terms of control burden of the following strategy, we hereafter reason in terms of per capita contact ratio (PCCR), denoted by c. This dimensionless number aims to quantify the average potentially infectious contacts an individual has per unit of time, relative to the pre-epidemic baseline (see Appendixes S2.2 and S2.3 for more details about how *c* is formally introduced in the model). This definition has the following consequences:

- the average PCCR before lock-down is 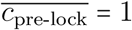,
- under the proportionate mixing assumption, the encounter rate at date *t* of two individuals belonging to age groups *i* and *j* respectively is proportional to *c_i_* (*t*) *c_j_* (*t*),
- the average PCCR during the lock-down is 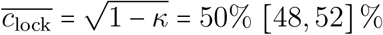, where *κ* is the lock-down control coefficient,
- the threshold PCCR value corresponding to a reduction of the reproduction number to exactly 1 is 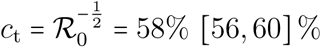.

By standardizing the control parameters of the model on a relative scale, the PCCR allows us to perform comparisons of control strategies that are independent from the estimated (past) or arbitrary (scenarios) values of both basic and temporal reproduction numbers. In addition, it provides an easy way to picture the control burden of each strategy. Indeed, the average PCCR 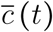 ranges from 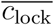 (the strongest control implemented so far) to 1 (the fully relaxed, ‘pre-lock-down’, situation). Between these two extremes, the remarkable value 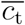 demarcates the threshold above which an epidemic rebound is made possible.

Notice that this formalism caputres both reduction in contact rate (e.g. working from home) and probability of transmission per contact (e.g. wearing a mask).

#### 3.4.1 Adaptive lock-down

The adaptive lock-down popularised by Ferguson et al. (2020) consists in triggering a lock-down (or possibly other broad restrictive measures) if an epidemic rebound is detected by a relevant indicator. This requires to carefully identify the threshold above which the lock-down is triggered. Following Ferguson et al. (2020), we use ICU admissions to quantify this threshold because they achieve a good balance between reflecting the epidemiological dynamics in the general population (the sampling of newly infected individuals is more homogeneous than with testing) and limiting the delay between the data and the state of the epidemics (we estimate this delay to be 2 weeks, while we estimate 4 weeks for mortality data).

Figure 6 shows that cumulative mortality is exponentially correlated with the daily ICU admission threshold, over the investigated range. As a consequence, there is no remarkable inflexion point that would justify a particular value, which would leave decision makers to coldly balance socio-economical costs (implied by low thresholds) with potentially saved lives (endangered by higher thresholds). Such a question is once again out of the scope of the present work.

**Figure 6:**
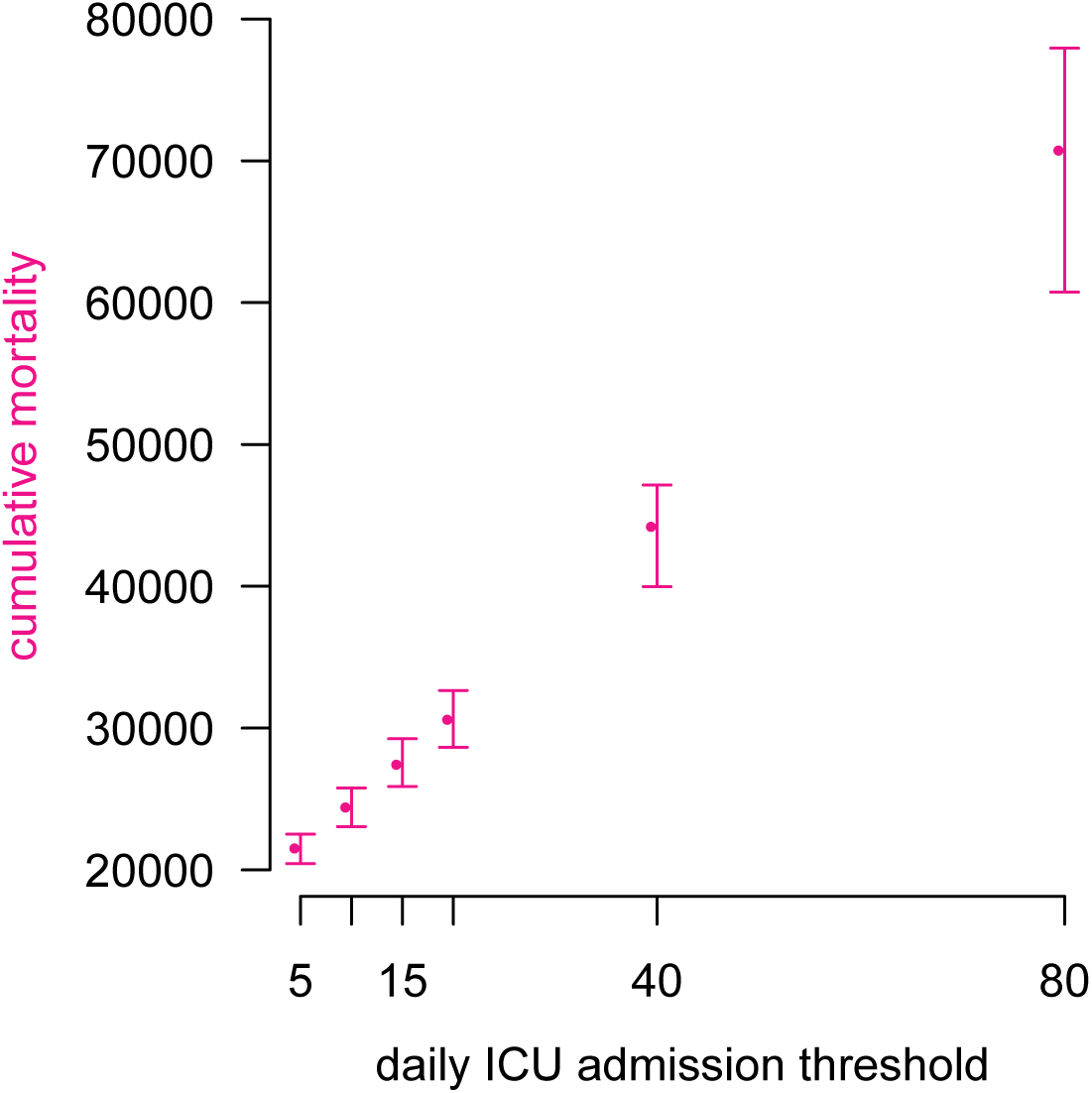
Adaptive lock-down threshold impact. Each dot represents a simulation of the model with adaptive lock-down implemented from May 12. The abscissa show the lock-down triggering threshold in terms of daily ICU admissions nationwide. The ordinate represents the median final death toll by the end of the year, along with their 95%-confidence intervals. Lock-down lifting threshold variation has negligible impact on the results (not shown here).

Figure 7 illustrates the chronicles produced by an adaptive lock-down strategy triggered by a threshold as low as 15 nationwide ICU admissions the same day. The epidemic can be efficiently controlled on the long run this way, at the cost of approximately 50 days of lock-down to only 12 days of release per cycle, that is ca. 20% of full release time, which is less than the proportion mentioned by Ferguson et al. (2020). This relaxation proportion could however be increased if moderate measures are still implemented between lock-down phases.

**Figure 7:**
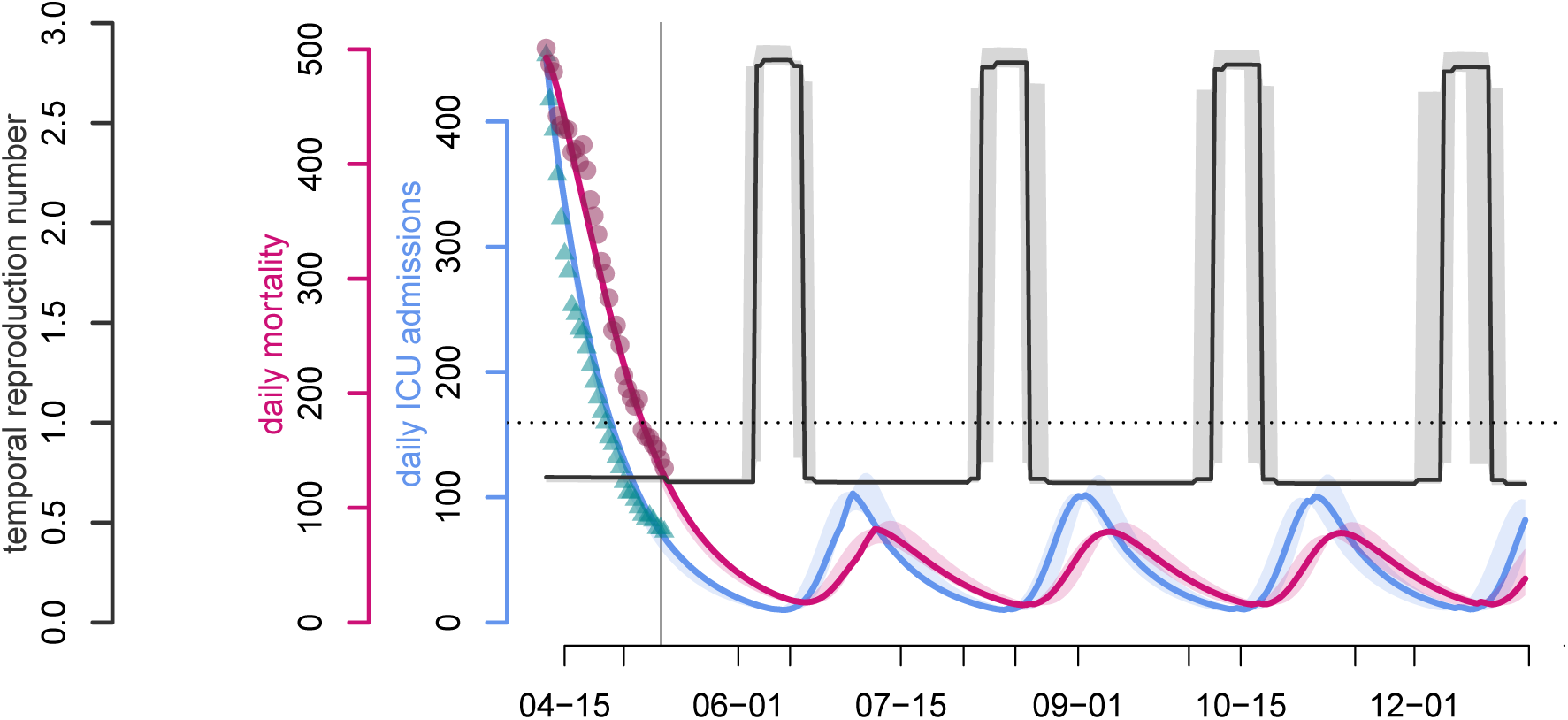
Adaptive lock-down waves. For the sake of clarity, only three parameter sets, chosen at random among the pool, illustrate here the dynamics generated by an adaptive lock-down strategy. In this scenario, the on and off thresholds are set to 15 daily ICU admissions nationwide. Symbols as in Fig.2.

#### 3.4.2 Periodic lock-down

By definition, an epidemic remains under control if 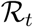 is kept below 1. If control measures alternate according to some periodic pattern, the instantaneous value of 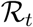 is of little interest and we consider instead its average value over one cycle of interventions, denoted by 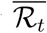. For the sake of simplicity, we consider a NPI that alternates between a hard phase (typically, a lock-down) and a relaxed (or release) phase, independently from any field indicator. Let us denote by *c*_r_ and *c*_h_ the average *per capita* contact ratio during the relaxed and hard phase respectively and *p*_r_ the time proportion of each NPI cycle allowed to the relaxed phase.

The epidemic is under control if

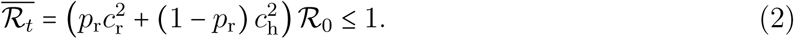

The NPI cycle can contain more than two phases, but one can always pool them into those favorable to social and economic life, represented by the couple (*c*_r_,*p*_r_), and those concentrating the necessary public health restrictions, namely the couple (*c*_h_, 1 − *p*_r_).

Assuming that hard phases have the same effect as the first lock-down, we can set 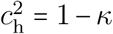 (see Appendix S2.3). Then, the maximum proportion of time allowed to the relaxed phase is given by

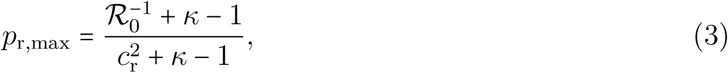

which is a decreasing function of *c*_r_. This expression captures the intuitive trade-off, shown in Figure S-8, which is that the lower the control in the relaxed phases, the shorter they can be. If there is no control in the relaxed phase (*c*_r_ = 1), then, based on our estimates for 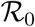 and *κ* (Table S-5), controlling the epidemics requires that hard control be implemented 88% of the time.

Here, we maximize the product between intensity and duration *(p*_r,max_*c*_r_) because it partly captures the absolute degree of freedom for a fixed period of time (i.e. try to be the less constrained as possible for the longest time period as possible). Noticing that 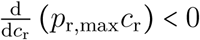 leads to the conclusion that the optimum is to seek for the lowest *per capita* contact ratio, which is obtained by setting equation 3 equal to 1. This yields 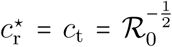 (which equals approximately 58% for 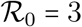). Under these conditions, there is no need for any harder phase.

Importantly, the simple product *p*_r,max_*c*_r_ does not need to be the objective quantity to maximize and parametrizations of 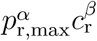, with *α, β >* 0, based on socio-economical arguments, should be investigated. However, this question is out of the scope of the present work.

Up to now we only investigated the proportion of the time spent in a relaxed or hard control phase. However, a 0.5 proportion can correspond to a 1 week / 2 week periodicity or to a 1 month / 2 month periodicity. In fact, the trade-off relation shown in Figure S-8 theoretically works for any time split where a proportion *p*_r,max_ is spent in a relaxed phase with per capita contact ratio *c*_r_ and a proportion 1 − *p*_r,max_) corresponds to a strong control phase where per capita contact ratio is restricted to *κ*. To be implemented, such periodic control should consider phases lasting at least several days. Furthermore, a periodicity greater than a month exposes populations to a greater risk during the relaxed phase.

Figure 8 suggests that the duration of the overall cycle has an effect up to 5,000 deaths, over the explored range. A weekly cycle yields the lowest death tolls, both in terms of median and CI. Furthermore, if the period lasts a month or less, a lower control of the epidemic in the relaxed phase yields the lowest cumulative mortality. Morecisely, these results shows that allowing 1 moderately relaxed (the average PCCR being equal to 80%) day per week within a prolonged lock-down has a better mortality outcome than the adaptive lock-down set with 15 daily ICU admission as a threshold (approx. 8,000 deaths less by the end of the year).

**Figure 8:**
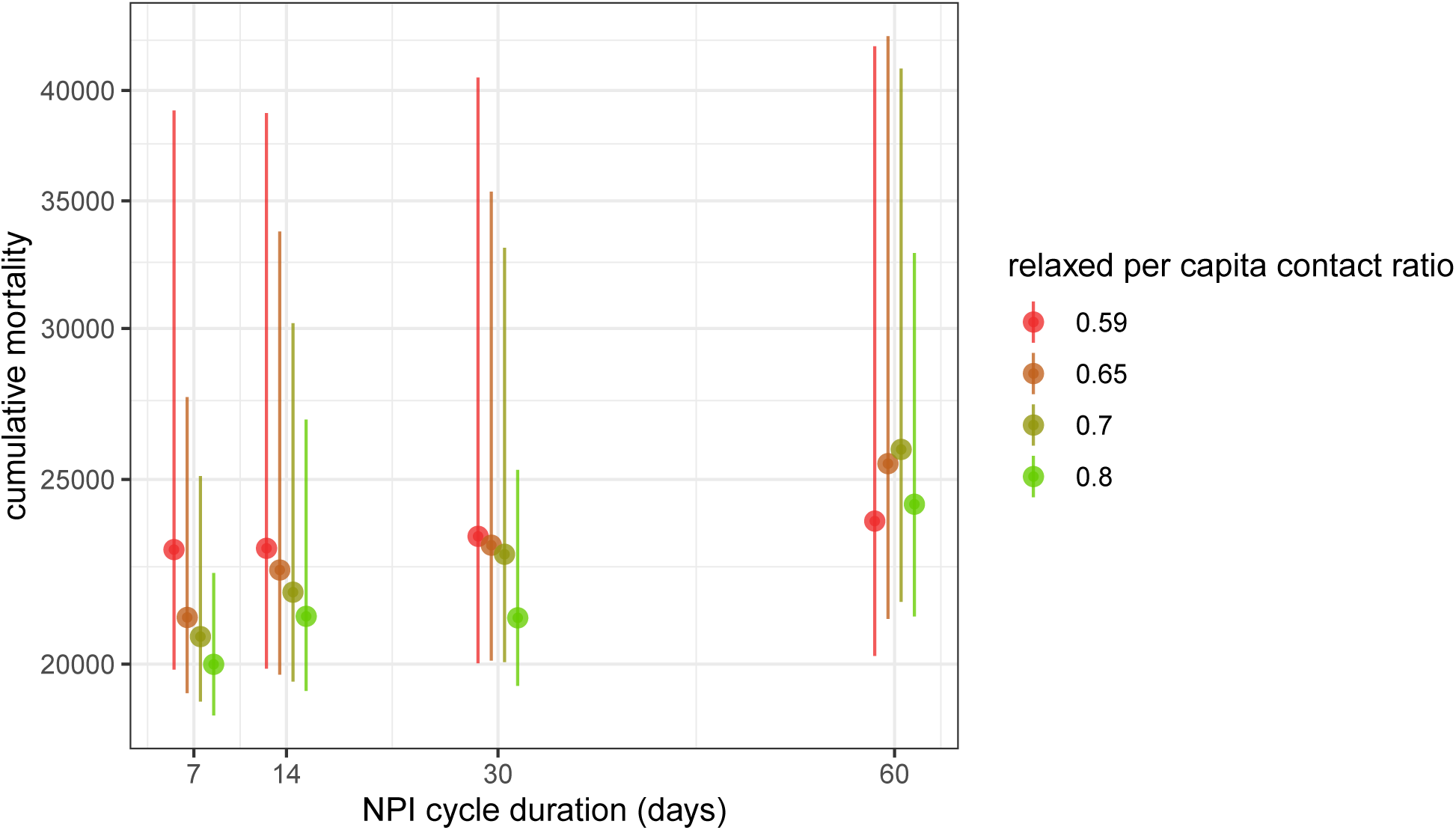
Effect of NPI cycle duration on cumulative mortality. We show the cumulative mortality by the end of the year 2020 if a periodic control is implemented from May 12. For each of the four NPI cycle durations, the model is run for four values for *c*_r_ shown in different colors, with a proportion of time satisfying eq. 3 (truncated to an integer number of days). For example, in the red scenario, *p*_r,max_ (0.59) = 88%, so for the weekly cycle the relaxed phase was set to floor (0.88 · 7) = 6 days.

#### 3.4.3 Age-differential control

Age-group targeted control measures are motivated by the age dependent risk profile (Verity et al., 2020a; Collaborative et al., 2020). However, this model could easily include other risk factors such as heart disease, diabetes, or obesity. One of the simplest strategies is to incite people at risk to stay at home. On the other side of the age pyramid, a finer age-specific strategy could be to close schools and universities. More generally, political and economical incentives to telework could also be considered as age-specific control measures.

We divided the population into three groups according to the cut-off ages of 25 and 65 years old. The first group is motivated by the control leverage represented by school and university closure, while the third is motivated by preventing viral circulation within the age group having the highest IFR. Each age group was assigned a fixed PCCR until the end of the year: 50, 58, 62, 65 and 75%. This led to 125 scenarios that are shown in Figure 9. The ratio of daily interaction between two individuals relative to the pre-epidemic baseline is the product of their individual contact ratios.

**Figure 9:**
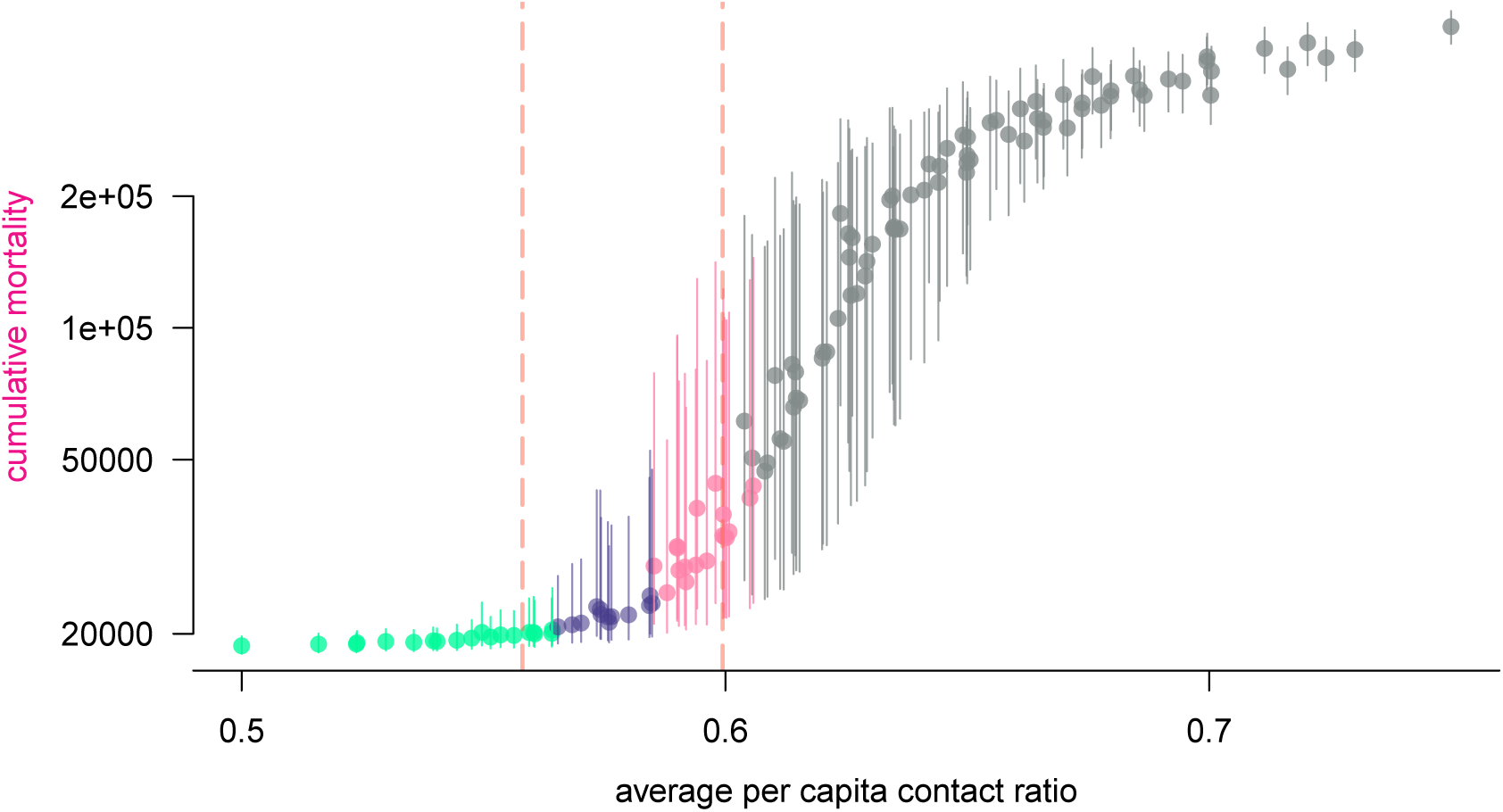
Effect of age-differential control interventions on cumulative mortality by the end of 2020. Each dot represents a simulation of the model with age-differential restrictions implemented after May 11. The abscissa corresponds to the demographic-weighted average in PCCR and the ordinates represent the median final death toll by the end of the year if no other solution is implemented (bars indicante 95%-CI). The color of the dots shows the temporal reproduction number by May 18: in green the epidemic is under control and the 97.5% quantile is less that 1, in purple the median is below 1 but the 97.5% quantile is above 1, in pink the median is above 1 but the 2.5% quantile is below 1, and in grey the epidemic is out of control, the 2.5% quantile is greater than 1. Vertical bars show the 95%-likelihood interval of the expected per capita contact ratio threshold above which the reproduction number is above 1, in a homogeneously-controlled population.

The results of the numerical investigation of 3-age group-differential controls are shown in Figure 9. This scatter plot shows that cumulative mortality is well explained by demographicaveraged PCCR. Therefore, the variance in contact ratio among age groups (not shown on the plot) has little impact on the long-term epidemic dynamics. This result suggests that there is no easy way of taking advantage of the strong correlation between COVID-19 complication risk and age, unless implementing an unrealistically restrictive quarantine for at-risk groups.

## 4 Discussion

COVID-19 is not an unusually lethal or contagious infectious disease. On the other hand, the large proportion of transmission attributable to individuals with few or no symptoms makes it redoubtably difficult to control compared to SARS (Fraser et al., 2004). In the absence of a massive and homogeneous screening effort, the reflection of the epidemic dynamics of COVID-19 is reduced to hospitalizations induced by complications, which are rare and occur on average two weeks after infection. The marginal, indirect and delayed nature of these events justifies the use of statistical analyses and mathematical modelling in the short-term epidemiological management of this unprecedented health crisis, that challenges all components of our societies. However, given the magnitude of the risks at stake, the diversity of fields involved, and the number of unknowns (such as climate effects, immunity duration), mathematical modelling predictions should be handled with caution in the decision process.

Informing decision making represents a challenge because most mathematical modelling in epidemiology relies on continuous time deterministic models. These offer a wide palette of analytical tools, but they also become less accurate on short time scales. Conversely, stochastic models, whether agent based or not, offer a much more precise picture of the early stages of the epidemics. However, they become less necessary once the outbreak threshold has been crossed and epidemic dynamics are essentially deterministic.

We therefore developed an original framework at the crossroads of individual-centered and compartmental modelling approaches, tailored to the COVID-19 natural history, which has two great advantages. First, its discrete time structure, shared with that of common epidemiological data, allows us to assume any distribution for model processes, therefore introducing what is known in the literature as memory effects (or ‘non-Markovian’ processes), related to the age of the infection. As a result, we obtain a much better fit than classical memory-less (or ‘Markovian’) models on intermediate timescales (weeks or months). Second, the deterministic nature of the model allows us to perform extremely fast simulations, especially compared to agent-based modelling that requires the drawing of millions of random numbers for a single simulation. The computational performance combined to the great parsimony, that still allows it to accurately fit the observed epidemiological dynamics, makes this model a relevant tool that can be easily transposed and deployed to other settings, countries or scales – even with data limited to those publicly available, as it is the case in the present work.

First, we used our model to infer three key parameters of the epidemics. Our 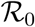 is comparable to that already computed in France based on their 95% confidence interval (Di Domenico et al., 2020; Roques et al., 2020; Salje et al., 2020; Hoertel et al., 2020). We also estimate the temporal reproduction number 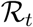 after the lock-down to 0.71, which is higher than that estimated by earlier studies (Salje et al., 2020; Hoertel et al., 2020), and the number of lives potentially saved (more than 50,000) compared to a lock-down implemented a week later.

Once the parameters estimated, we investigated potential scenarios for control, based on the assumption that the major characteristics of the epidemics will not change, meaning for instance that a seasonal effect would have a limited impact on transmission dynamics. Our results suggest that, in the case of a rise of the reproduction number above the unit threshold value, a reinforcement needs to be implemented within 5 weeks after lock-down lifting to prevent a massive second epidemic peak.

Then, we explore two types of cycling strategies. The first strategy, known as adaptive lockdown and popularised as ‘stop-and-go’, consists in alternating strict lock-down and absence of control (Ferguson et al., 2020), the shift between the two being based on the incidence of daily patient admission in ICU. We show that this requires a low threshold without a particular value standing out. The second strategy explored is the fixed-period alternation between lockdown and relaxed phases. Our work highlights that, for the adaptive lock-down to perform better than the latter, both in terms of casualties and time spent under relaxed control, part of the restrictions have to be maintained between lock-downs. Besides, we investigated age-differential control strategies and show that there is little public health benefit to be gained from the variance of control restrictions between age groups, unless implementing measures even more restrictive than the past lock-down. However, synergy between age differential and periodic control strategies is left to be explored.

The model makes several strong assumptions. First, there is no spatial structure. This limitation can become strong if the epidemic grows in size to infect a large proportion of the population. Second, there is no specification of the public health control measures implemented: all the options (quarantine of confirmed cases, adoption of barrier measures, social distancing: closing of schools and universities, banning of gatherings, etc.) are combined to reduce the contact rate. We also neglected fomite transmission (see Ferretti et al. (2020) for an example) and assumed perfect and lifelong immunity against reinfection due to currently insufficient data on immunity. One simplifying assumption we made is that mortality probabilities do not vary over time, whereas in practice hospital saturation could affect mortality, whether related to COVID-19 or not.

In terms of outlook, this work lays the foundation for an online application soon to be released, as an update of the earlier version COVIDSIM-FR (ETE Modelling Team, 2020a). Next challenges include taking to account for possible changes in parameter values with time, mainly detecting and estimating seasonal effects. If these are of significant impact, model fitting could be adapted, either by estimating parameters within defined time windows or by left-censoring the data as time goes by. Lastly, the NPI analysis exposed in this work sets the ground for the exploration of finely tuned public health measures accounting for spatial heterogeneity and combining advantages of adaptive, periodic and group-differential modalities, with the aim of avoiding an epidemic rebound while minimizing its population burden.

## Data Availability

All the data here used are publicly available and their references indicated.

## Acknowledgements

This work is based on a report and online shiny application published online on Apr 6, 2020 http://bioinfo-shiny.ird.fr:3838/COVIDSIM-FR (ETE Modelling Team, 2020a).

We acknowledge support from the University of Montpellier, the CNRS and the IRD.

## Authors’ Contributions

Model design and analysis performation: MTS

Manuscript redaction: MTS, SA, YM

Data curation: BR, BE

Computational support and sofware development: BR

Mathematical support: RDD, CS

Project conception and validation: all

## Appendix

**Table of Contents**

**S1 Mathematical notations 2**

**S2 Model details 5**

S2.1 Recurrence relation system 5

S2.2 Force of infection 5

S2.3 Lock-down effect 7

S2.4 Times series 9

S2.5 Criticality-related probabilities 9

S2.6 Waiting times 13

S2.7 Fitting and estimation procedure 15

S2.8 Derived outputs 16

S2.9 Continuous time model 18

**S3 Supplementary Results 19**

S3.1 Maximum likelihood parameter estimates 19

S3.2 Data right censoring and parameter inference 20

S3.3 Lock-down implementation date and cumulative mortality 23

S3.4 Examples of post-May 11 scenarios 24

S3.5 Time vs intensity trade-off of the relaxed phase in the periodic lock-down strategy 25

### S1 Mathematical notations

Summary table with all the notations used in the study.

**Table S-1:**
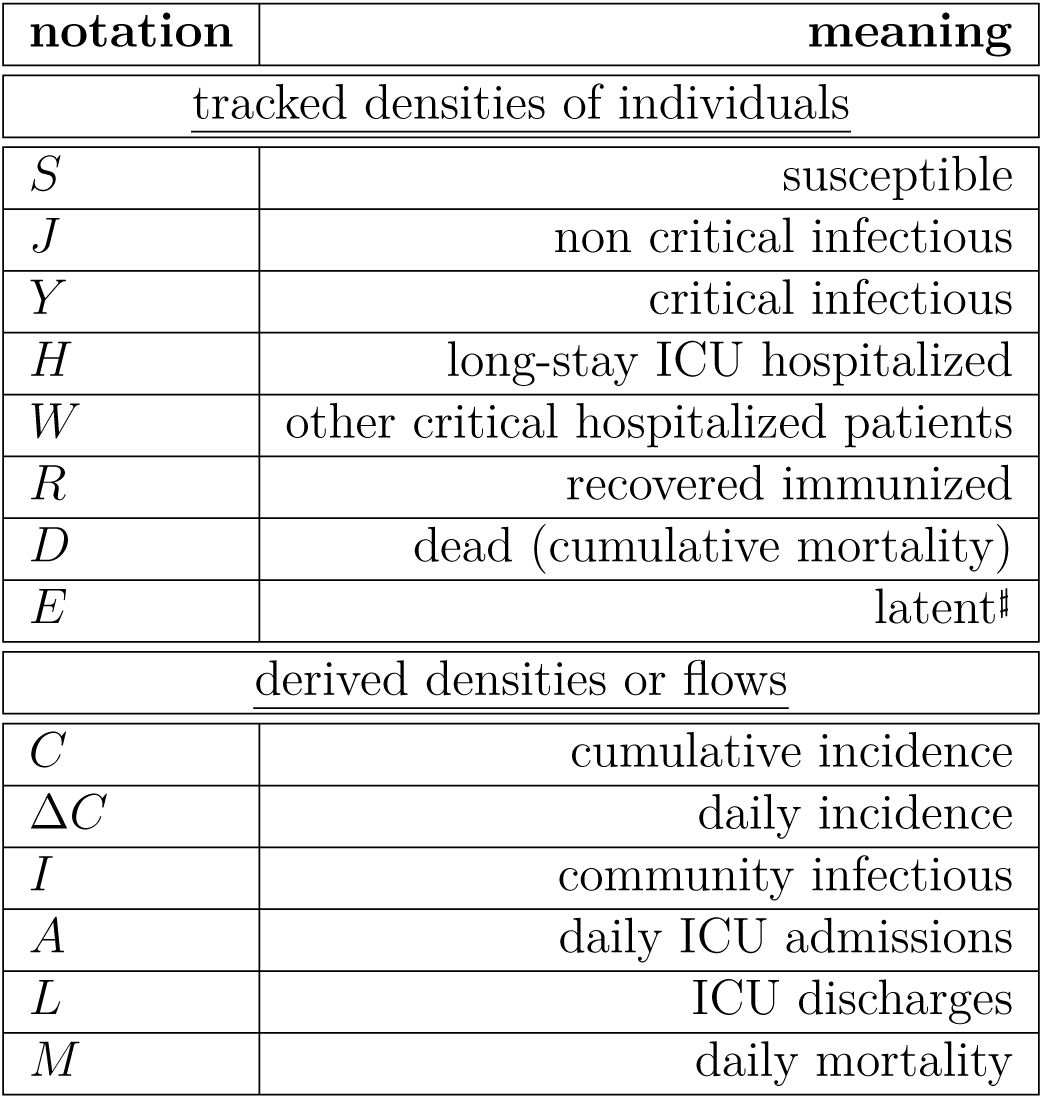
Density related notations. #: only applies to the Markovian continuous-time model.

| notation | meaning |
| --- | --- |
| <u>tracked densities of individuals</u> |  |
| $S$ | susceptible |
| $J$ | non critical infectious |
| $Y$ | critical infectious |
| $H$ | long-stay ICU hospitalized |
| $W$ | other critical hospitalized patients |
| $R$ | recovered immunized |
| $D$ | dead (cumulative mortality) |
| $E$ | latent<sup>#</sup> |
| <u>derived densities or flows</u> |  |
| $C$ | cumulative incidence |
| $\Delta C$ | daily incidence |
| $I$ | community infectious |
| $A$ | daily ICU admissions |
| $L$ | ICU discharges |
| $M$ | daily mortality |

**Table S-2:**
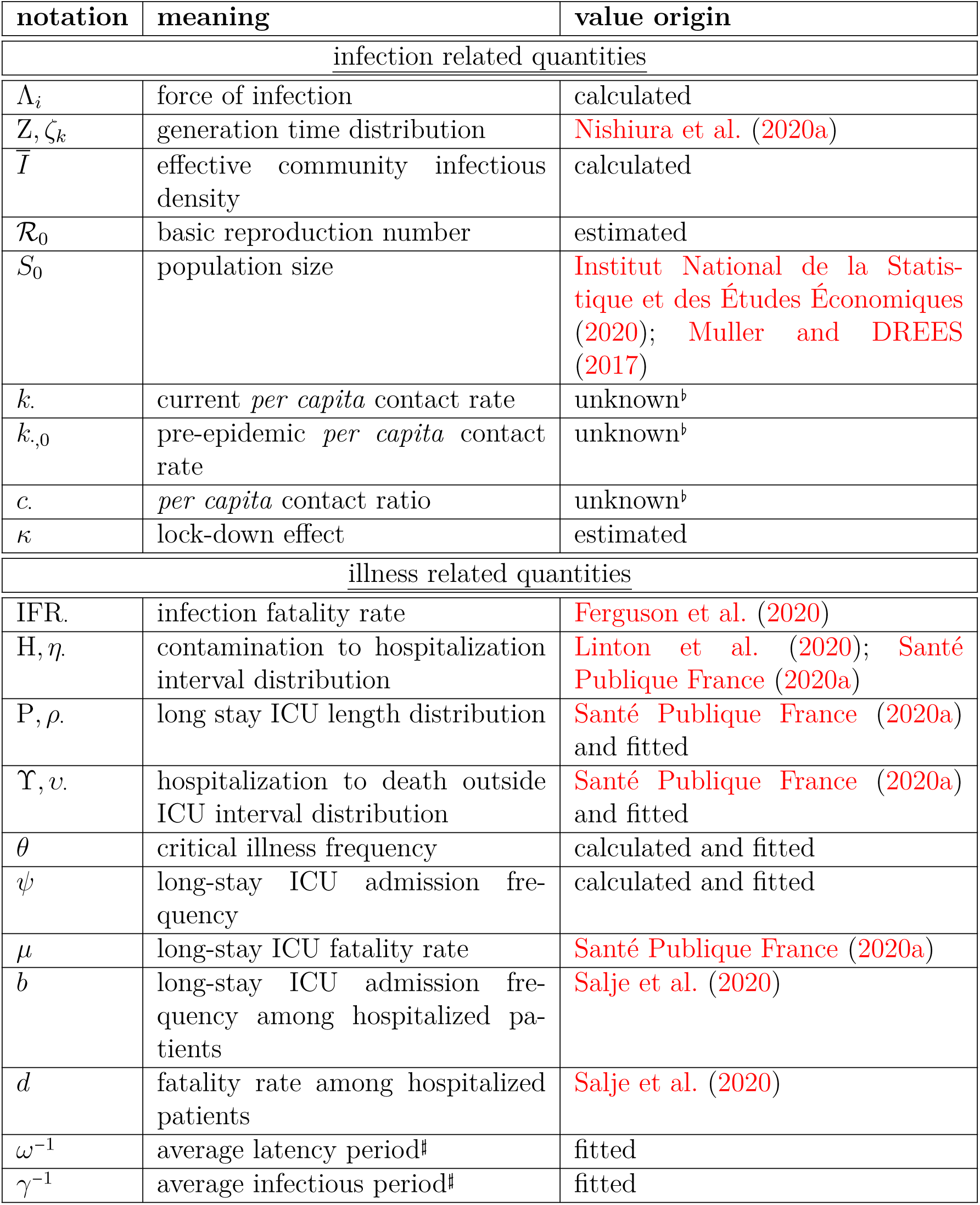
Main parameter notations. #: only applies to the Markovian continuous-time model. *b* the calculation of these values are bypassed by the estimation of *κ*, as shown in S2.3.

**Table S-3:**
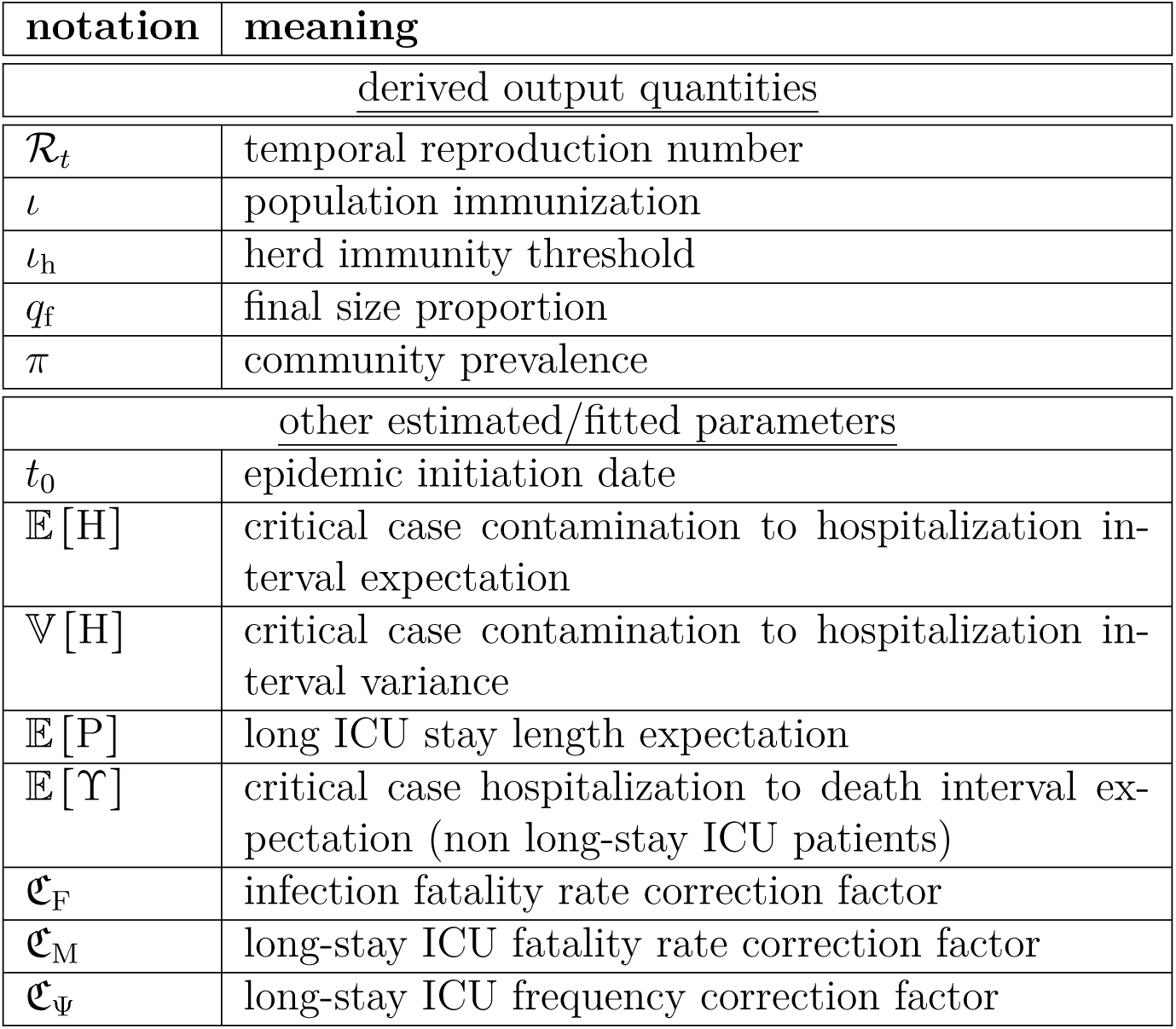
Output-related quantities and fitted accessory parameters.

**Table S-4:** Other generic notations.

| notation | meaning | value origin |
| --- | --- | --- |
| <u>other notations</u> |  |  |
| $\cdot_i$ | age group (average, density...) | calculated |
| $\tilde{\cdot}$ | moving-averaged data | Santé Publique France (2020b) |
| $\hat{\cdot}$ | adjusted parameter | fitted |
| $\mathfrak{f}.$ | demographic frequency | Institut National de la Statistique et des Études Économiques (2020); Muller and DREES (2017) |

### S2 Model details

#### S2.1 Recurrence relation system

Instead of classical ordinary differential equations (ODE), the dynamics of the model satisfy a system of recurrence relations (one could as well write as finite difference equations (FDE)), which is detailed below. For the sake of simplicity, we omit time dependence in the notations. Instead, *X_i_* denotes a density at a given time *t* and 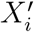 the density at time *t* + 1. The equations are formally identical for all age groups *i*. Between-group dynamics are coupled through the forces of infection Λ. defined in the next subsection.(S-1)

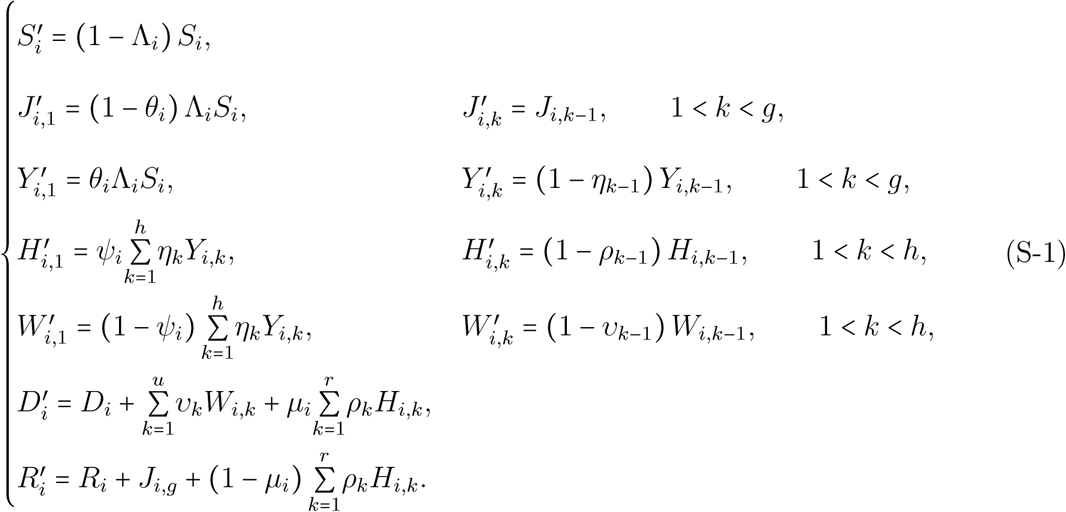

#### S2.2 Force of infection

As indicated in the main text, our goal is to have the force of infection function follow the well-known Michaelis-Menten (or Holling type II physiological response) function of the form 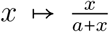. However, we cannot use the prevalence as the argument of Λ*_i_* here because all infected individuals, whether they are critically ill (*Y*) or not (*J*), do not contribute equally to transmission events. This heterogeneity in contagiosity originates from differences in infection ages (individuals contaminated 6 days earlier are more contagious than those 10 days earlier) and in contact rates.

To address this issue, we introduce the effective infectious density 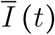,

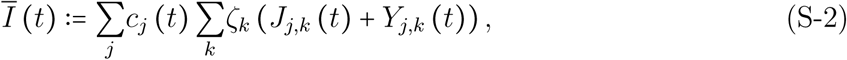

which is the sum over all infected community compartments weighted by both the generation time *ζ_k_*, which is the time between the infection of an ‘infector’ and the infection of his or her ‘infectee’, and the per-capita contact ratio *c_i_* (*t*). The latter is defined as the current contact rate per-capita of individuals of age group *i* (*k_i_* (*t*)) relative to their pre-epidemic baseline contact rate (*k_i_*,_0_),

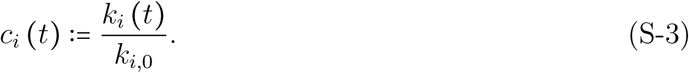

With 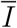 kept constant, Λ*_i_* is expected to display a Michaelis-Menten behavior (i.e. positive initial slope, increasing and upper-bounded) with respect to the current per-capita rate as well. Consequently, the force of infection should satisfy

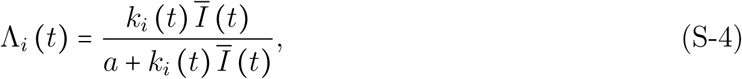

as shown in Fig.S-1.

We then derive the expression of *a* using known parameters. To do so, we consider the probability for a given susceptible individual to be part of the first generation of cases, that is, to have been infected by the index case. Let us denote Λ*_i_*_,index_ *(t)* the probability of being infected by the index case on day *t*. By definition of the basic reproduction number, the index case infects 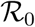 secondary cases, on average, by the end of its contagious period. Under the mean-field approximation, the probability of being part of these secondary cases is simply 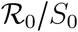 (which is an extremely rare event). Summing over all possible days, we therefore have

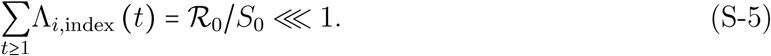

For simplicity, we make the following three assumptions:

- the index case is not critically ill (less than 5% of cases are),
- the index case belongs to the same focal age group *i* (More generally, one should average the per-capita contact rate of the index case weighted by the (unknown) probability of her or him belonging to each age group. This uncertainty was circumvented in the numerical inferences and simulations by pooling all individuals within a single age group, the parameters of which were weighted according to demography),
- the index case has infected all his or her secondary cases before public health measures are implemented.

It follows from these assumptions that if we set the contamination day of the index case to *t =* 0, the effective infectious density 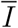 restricted to the index case can simply be written as

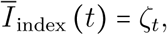

(note that *k_i_* (*t*) = *k_i_*,_0_ over the considered period of time, hence *c_i_* (*t*) = 1 and all densities are equal to 0 except *J_i,t_ =* 1).

Applying these results to equation S-4, the daily probability of infection by the index case therefore becomes

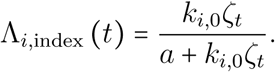

Now, from the magnitude comparison (S-5), we have 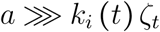, and hence

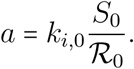

In the limit of low prevalence (or low contact rates), the mass action law is recovered. Using equation (S-5) and the fact that the sequence (*ζ_t_*)_t≥1_ being a mass function, it sums up to 1, we get

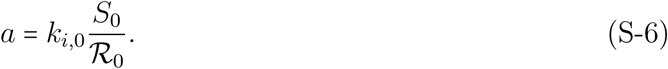

Combining equations (S-4), (S-3) and (S-6) finally leads to the generic expression of the force of infection:

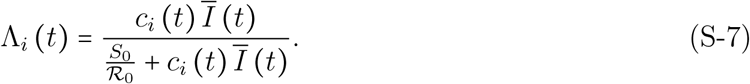

This is the equation indicated in the main text.

#### S2.3 Lock-down effect

In the special case where strong public health control measures such as lock-down are being implemented, all individuals may exhibit similar per capita contact rates, *k_i_* (*t*) = *k*_lock_. From (S-4), it is then possible to express the force of infection in the following way:

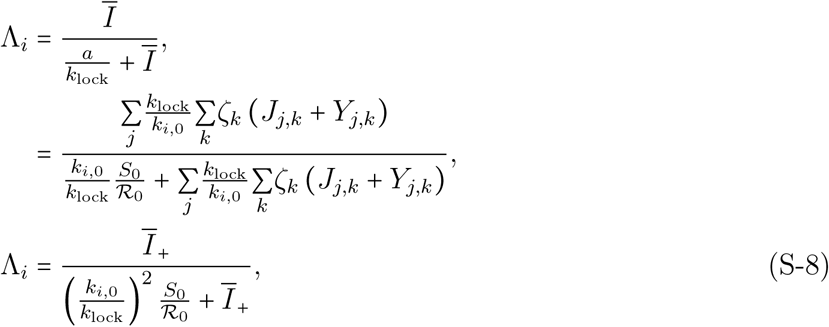

where 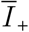 is the effective infectious density as if it were calculated in absence of health measures (i.e. with *c_i_* = 1). In equation (S-8), one can interpret the quantity 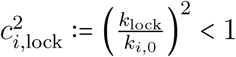 as a factor lowering the basic reproduction number 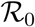. The lock-down effect, defined as the reduction of 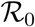 due to this measure, can be calculated by averaging *c_i_*_,lock_ over age groups (according to demography). Hence,

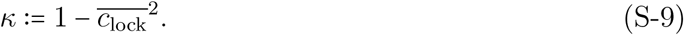

**Figure S-1:**
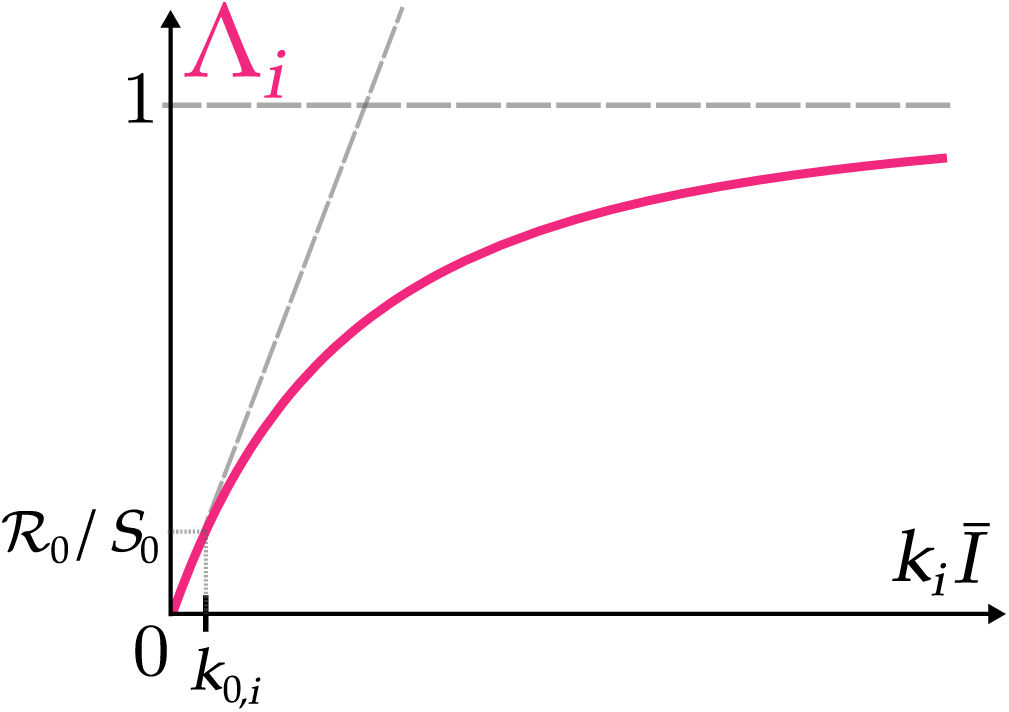
Daily force of infection as a function of the product of daily contact rate and effective infectious density. Λ*_i_* is the probability for one susceptible individual from group *i* to be infected a given day. The initial non-zero slope comes from the law of mass action implied by the mean-field approximation. The derivation of Λ*_i_* is based on the remarkable coordinates shown near the origin of the graph (not to scale). When the effective infectious density is equal to 1 (i.e. as if the generation time distribution were concentrated in a single day) and the contact rate is that in absence of any health measure (denoted by *k*_0,_*_i_*), then, by definition of the basic reproduction number, Λ*_i_* equals 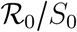.

#### S2.4 Times series

The largest and most reliable nationwide data for the COVID-19 epidemic in France is that of daily COVID-related death toll in hospitals, communicated daily since Feb 16 2020 by the national public health agency (Santé Publique France). This is why our model neglects COVID-related deaths occurring outside hospitals. In particular, we removed nursing homes (or EHPAD) from calculations.

Starting from Mar 18 2020, two additional time series are communicated: daily admissions in intensive care units (ICU) and current ICU occupied beds. While the former capture the dynamics from contamination to ICU admission, the latter captures moreover the kinetics of ICU stay.

These time series are altered by week-ends and bank days: e.g. death tolls are notably lower on Sundays than previous days, while it increases the next Mondays. It has even been suggested that reporting delays propagate also to Tuesdays. In order to smooth these artifactual weekly oscillations, a right-shifted 7-days moving average was performed over all time series prior to analysis. We will refer to these smoothed datasets as 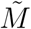 for daily hospital mortality, *Ã* for daily ICU admissions and 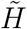 for current ICU occupied beds. Since 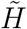 contains part of the cumulative information of *Ã*, we also considered 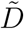, the cumulative counterpart of 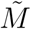 to equilibrate the first step of the fitting procedure (see below).

#### S2.5 Criticality-related probabilities

Symptom severity of COVID-19 is increasingly classified into mild, moderate, severe and critical. Because we rely on hospital mortality and ICU flow data, we focus on critical cases, i.e. the ones concerned by intensive care and COVID fatality. In the absence of detailed large-scale hospitalization data, we made the following assumptions:

- non-critical cases are not admitted into ICU (even though some do need hospitalization),
- all critical cases need hospitalization, and only survive if they go through intensive care.

For medical reasons not addressed here, in France not all critical cases are admitted soon enough into ICU. Part of them die in non-intensive care wards, while others die shortly after entering the ICU, therefore not contributing to ICU bed occupancy. We therefore need to estimate two key criticality-related probabilities, namely *θ_i_*, the proportion of critical cases within age group *i*, and *ψ_i_*, the proportion of critical cases in age group *i* that contribute to ICU bed occupancy (i.e. their stay in the ward exceeds one day). Because these probabilities differ among age classes and improper averaging could lead to substantial bias (see below), we first need to make calculations focused on the smallest age stratification unit (usually a decade). In the following, *θ* (*a*) and *ψ* (*a*) are the age-specific critical case frequency and long-stay admission given critical illness respectively. In addition, ℙ*_a_* [X_1_|X_2_] reads as the probability of event X_1_ given X_2_ has occurred for an individual of age *a*.

Let us consider the four events needed to derive *θ* (*a*) and *ψ* (*a*) from available data:

- I, being infected by SARS-CoV-2,
- U, being hospitalized,
- B, occupying an ICU bed for more than a day,
- D, dying at the hospital from COVID-19.

The interplay between these events is formally depicted by the tree diagram in Fig.S-2.

**Figure S-2:**
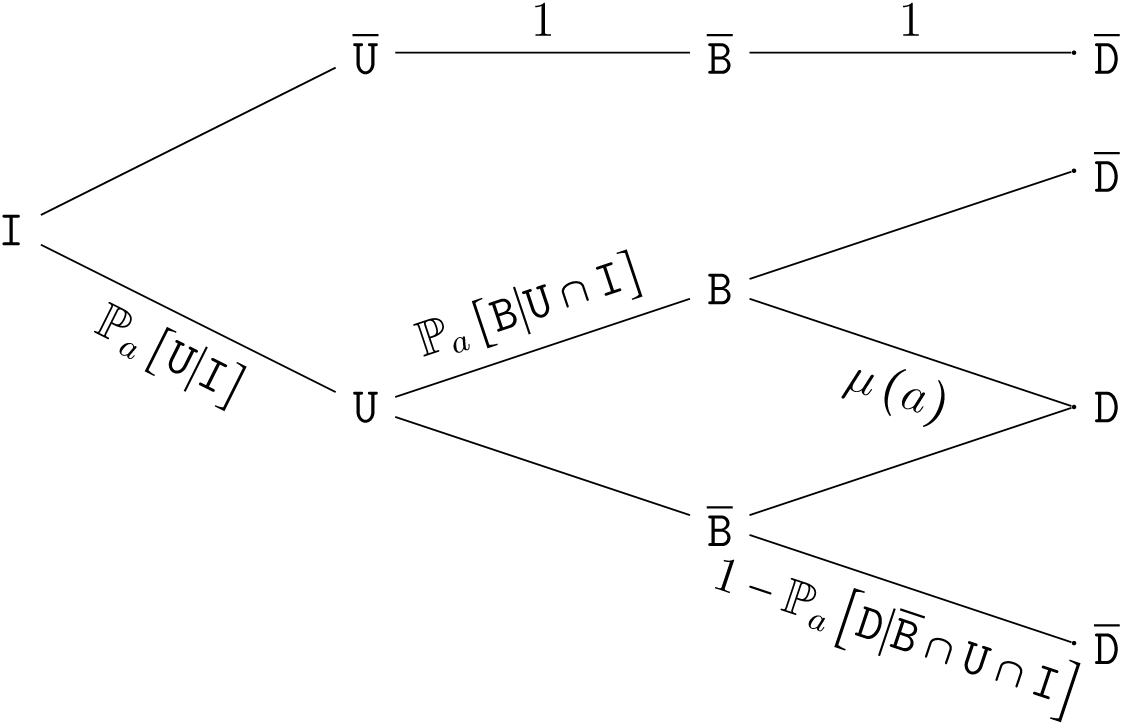
Tree diagram of critical COVID-19 related events. A fraction ℙ*_a_* [U|I] of infected (I) are hospitalized (U). Among these, a proportion ℙ*_a_* [B|U ∩ I] are admitted into ICU for a stay longer than a day B. The fatality ratio (D) equals *μ* (*a*) for these patients and 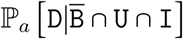 for the others.

We need to find two independent equations involving probabilities related to these events for which age-stratified data is known in order to solve *θ* (*a*) and *ψ* (*a*). First, we need to identify such data:

- ℙ*_a_* [D|I] is the proportion of deaths among COVID-19 infected individuals, better known as the Infection Fatality Rate (IFR), which has been calculated using the Diamond Princess data by Verity et al. and corrected for non-uniform attack rate by Ferguson et al. and will be denoted by IFR (*a*) hereafter,
- ℙ*_a_* [D|B ∩ U ∩ I] is the proportion of deaths among COVID+ ICU hospitalized patients; this age stratified data has been communicated by Santé Publique France as a weekly epidemic report on May 7 2020 (Santé Publique France, 2020a), and will be denoted by *μ*(*a*) hereafter
- ℙ*_a_* [B|U ∩ I] and ℙ*_a_* [D|U ∩ I] are respectively the proportions of hospitalized patients (whether they are critical or not) admitted in ICU, and those that die (whether in ICU or not), and hereafter denoted by *b*(*a*) and *d*(*a*). These data come from the SI-VIC database and made available by Salje et al. (2020). (Tables S1 and S2).

A first equation comes by noticing that the probability for an infected individual to die from COVID-19 is the probability of developing critical illness if infected (namely *θ*(*a*)) times the proportion of deaths among critical cases. The latter is the sum of critical cases that die in ICU after a stay longer than one day (*μ*(*a*) *ψ*(*a*)) and the critical cases that are not lengthily admitted in ICU and cannot be saved (1 − *ψ*(*a*)):

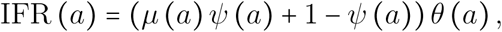

hence

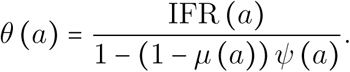

Now, to find *ψ*(*a*), let us notice that the proportion of deaths not occurring in ICU can be expressed as

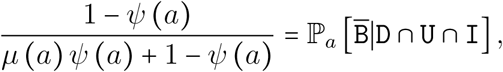

hence

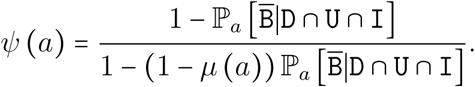

The unknown probability can be calculated as follows

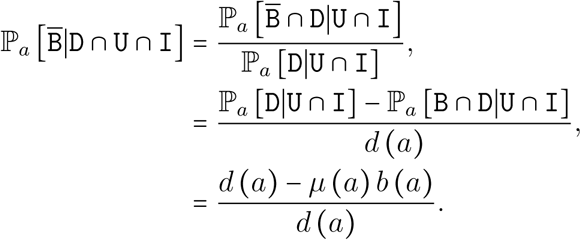

After elementary calculations, we finally get

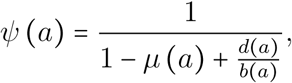

hence both *θ*(*a*) and *ψ*(*a*) can be calculated from age-stratified available data.

The mean proportion of critical cases among a specific age group, *θ_i_*, is simply the demographic-weighted average of *θ*(*a*) over the considered ages, as infection samples uniformly the susceptible compartment (NB: the IFR stratified data used here from (Ferguson et al., 2020) is already corrected for non-uniform attack rate). However, as the probability of the next events related to critical illness are not homogeneous with respect to age, the age group averages *ψ_i_* and *μ_i_* cannot be weighted directly with relative demographic age frequencies. Instead, they must be calculated by taking into account that ages with higher *ψ*(*a*) and then *μ*(*a*) will be over-sampled in *Y_i_* → *H_i_* and *H_i_* → *D_i_* transitions respectively.

To account for this bias and as well to allow adjusting the parameters to both the model (for *ψ*(*a*) and *μ*(*a*)) and the French epidemic (for the IFR), we introduce a series of corrections detailed hereafter in the calculation of the parameters used to run the model.

First, we account for the fact that the ICU fatality rate might mix both short and long-stay patients, while our model splits these two flows. The corrected age-specific long-stay ICU fatality rate will be denoted by 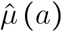 and calculated as the product of the corresponding data and a correcting factor denoted by 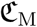, i.e. 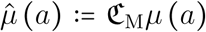. Likewise, the age-specific IFR (which was not estimated from French data) will be corrected as 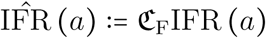. Now, equations S2.5 and S2.5 rewrite as

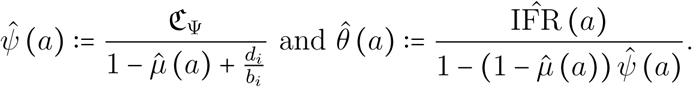

Note that there is no need for a fourth correction factor as 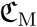, 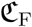, and 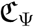 already capture possible corrections for 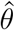. These corrections are yet of unknown values but will be estimated by the fitting procedure detailed below. As the corrected values are still probabilities, the correction factors are necessarily upper bounded, respectively by (max*_a_μ* (*a*))^−1^, (max_a_IFR (*a*))^−1^, and (max*_a_ψ* (*a*))^−1^.

The last step consists in age-group averaging as mentioned above. Let us denote by 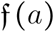 the frequency of individuals of age *a* in the French metropolitan population (after having removed the ca 730,000 individuals living in nursing homes). We call the i-group relative frequency of age *a* as the standardized age frequency 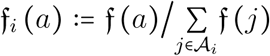, where 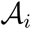 is the set of ages *i* belonging to age group *i*. As previously implied, the frequency of critical cases in age group *i* is the straightforward demographic weighted average

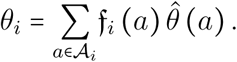

The frequency of long-stay ICU patients among hospitalized critical cases in age group *i* is then weighted by both the relative age frequencies and the ratio of critical illness probability to the group average, i.e.

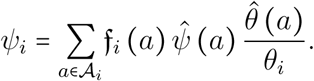

Finally, as the last event to occur, the average fatality ratio for long-stay ICU patients belonging to group *i* must be corrected by the ratio of the product of the two previous frequencies relative to the group average *ϕ_i_*, i.e.

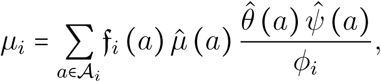

where 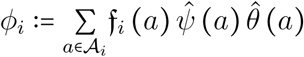.

#### S2.6 Waiting times

Four time distributions underlie the dynamics: the generation time (or index-contamination-to-secondary-contamination interval), the contamination-to-hospitalization interval of critical cases, the ICU length of stay and the hospitalization-to-outside-ICU death interval of critical cases. Each of these events can be seen as a random waiting time variable, denoted by Z° (capital zeta), H° (capital eta), P° (capital rho) and ϒ° (capital upsilon) respectively.

Initially, all four random variables are assumed to follow Weibull distributions, with shape parameters greater than one. Such distributions are widely used in the biomedical literature, along with Gamma and Lognormal distributions, for fitting the probability density function (PDF) of ageing processes, for which the probability for the focal event to occur increases with lapsed time (Bolker, 2008). Preliminary exploratory fittings of the model to daily mortality data indicated that maximum likelihood estimates of the shape parameter of P° and ϒ° were close to unity. Because the computational procedures used for maximum likelihood estimation misbehave in the vicinity of parameter range boundaries, the shape parameter of these two distributions was set to 1, turning them into exponential distributions by fitting (the exponential distribution being a special case of Weibull distributions), though not by assumption.

Weibull distributions have a right-unbounded support [0, ∞), which means that true distributions require truncation for obvious computational reasons. Let us introduce the generic notations Ξ ≡ Z, H, P, ϒ and *x* ≡ *g,h,r,u*. We construct the right-truncated analogous distributions by setting the finite upper boundary of their support 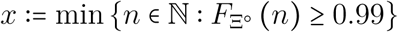, i.e. the upper-integer-rounded 99%-quantile of the original distribution Ξ°, where *F*. denotes the cumulative distribution function (CDF). The truncated distributions Ξ are therefore such that their CDF satisfy 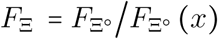, and having as their supports *F*_Ξ_ (Ω) *=* [0; *x*].

Dynamics unfold in discrete time in our model, which means these continuous distributions need to be discretized into sequences to be implemetend into the framework. The generation time sequence is straigtforwardly defined as *ζ_k_*: = *F*_Z_ (*k*) − *F*_Z_ (*k* − 1) for 1 ≤ *k* ≤ *g*. Indeed, transmission events do not affect the progression of the infector within its compartment. However, the three other sequences of parameters, *ξ_k_* ≡ *η_k_,ρ_k_,u_k_*, represent the proportion of individuals that leave the compartment *k* days after having entered it. These parameters need to capture the probability that the corresponding event occurs on day *k* but they also need to be standardized by the probability of not having left the compartment by that day. They are therefore calculated as

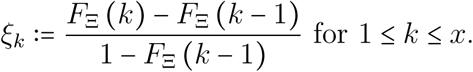

#### S2.7 Fitting and estimation procedure

The fitting procedure was performed using the mle2 routine from the bbmle package (Bolker et al., 2020) implemented in R (R Core Team, 2020). Starting from the initial parameter values v_0_, an ordinary least square optimum **v**_1_ was found by minimizing the euclidean distance to *Ã*, 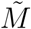, 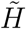 and 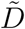 simultaneously, thus accounting for all events, from contamination to death or recovery. Having great confidence in both ICU admissions and current ICU occupancy is especially valuable for forecasting hospital needs, while mortality predictions cannot be ignored by decision makers. This first step is only used to locate the closest parameter region from **v**_0_ where likelihoods further calculated might reach their maximum value.

Maximum likelihood estimates (MLE) and 95%-likelihood intervals (LI) were calculated using the same routine. We assume observed data to be Gaussian-noised realizations of the model prediction, then considering each daily count to be distributed as 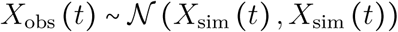, where *X*_sim_ is the simulated daily count. The choice of the distribution is supported by the large numbers involved and the Poissonian nature of count processes (NB: pre-lock-down mortality data was ignored for the central limit to apply). Contrary to the first step, only one time series was used for each estimation. MLE and LI of 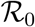, *t*_0_, 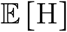, 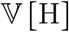 and *κ* were estimated with respect to *Ã*, while the other parameter values were set as in **v**_0_. The resulting MLE of the free parameters replaced the corresponding values in **v**_0_, providing vector **v**_1_. This new parameter set served as the starting point to estimate the MLE and LI for 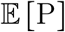, 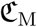 and 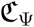 with respect to 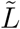, the derived time series of daily ICU discharges (calculated as the daily ICU admission minus the daily difference in ICU bed occupancy). The resulting parameter set **v**2 then initiated the estimation of the remaining parameters, 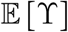 and 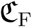, with respect to 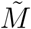, giving vector **v**_3_.

Using the fact that the maximum likelihood estimators are asymptotically normally distributed, we used the range of the LI as proxies for the standard deviation of the marginal distribution of each MLE. Then, we randomly drew parameter sets in a multivariate Gaussian distribution with mean **v**_3_ and diagonal variance-covariance matrix. For each of them, we calculated their likelihood with respect to 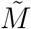 and kept only those whose likelihood was not significantly different from that of **v**_3_, according to Wilk’s theorem (Wilks, 1938). Sampling stopped once 10^3^ draws have satisfied the condition. Importantly, we considered all the retained parameters equivalent from the likelihood point of view, i.e. they are assumed to represent equally likely versions of adjusted parameter sets. Their diversity thus allows to account for uncertainty in the real parameter values.

For any further analysis of the model, system S-1 was run independently with each of the 10^3^ parameter sets. The confidence intervals of simulated tracked densities (*S*(*t*), *J*(*t*)…) as well as any derived quantity 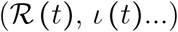 were then simply calculated for each time point as the unweighted 2.5% and 97.5% sample quantiles of the 10^3^ outputs at the given time point. The central estimations correspond to the median value of these distributions.

#### S2.8 Derived outputs

Tracked densities are the number of individuals in each clinical-epidemiological compartment *X_i,k_*, the dynamics of which satisfy S-1 and are thus directly provided by numerical iteration of the recurrence relations. However, several quantities of interest require additional calculations, hereafter exposed.

Let us first introduce two notations for the sake of concision. Tracked densities without indices will refer to sum over all groups, and, for multiple-days compartments, over all possible days of progression as well: 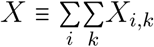 (e.g. *J*(*t*) represents all individuals belonging to *J_i,k_* for all groups *i* and all ages of infection *k*). The daily difference *∆X(t) ≡ X*(*t*) − *X*(*t* − 1) is straightforwardly used to extract the instantaneous dynamics from a cumulative time series.

The following three time series are crucial as they are used for likelihood calculations with respect to their data counterpart:

- *M*(*t*):= *∆D*(*t*) is the daily mortality,
- 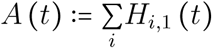 is the daily ICU admissions number,
- *L*(*t*):= *A*(*t*) − *∆H*(*t*) is the daily ICU discharge number.

The next times series are intermediate calculations required for further key quantities.

- *I*(*t*):= *J* + *Y* is the community infectious density and represents all not hospitalized infected individuals, which can be used to estimate the expected proportion of COVID PCR+ in the general population,
- 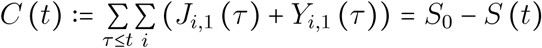 and ∆*C*(*t*) are the cumulative and instantaneous incidence respectively.

The latter is used for the calculation of the most scrutinized indicator in epidemic monitoring, namely the temporal (or effective) reproduction number, 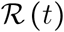, which we calculate here following Wallinga & Lipsitch, at the time of the infectees’ contamination:

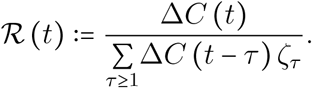

In this work, (population) immunization 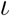 refers to the proportion of individuals that have been infected by SARS-CoV-2. We assume waning immunity is negligible at this timescale. Its calculation is simply

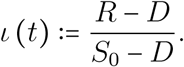

In absence of waning immunity from the host and antigenic drift from the virus, and assuming public health measures are fully relaxed, further epidemic can only be passively prevented if the herd immunity threshold is reached, i.e. 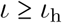 where

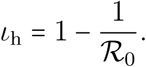

A classical result from Kermack & McKendrick is that if the epidemic has started spreading, the final cumulative relative incidence will not stop at the herd immunity threshold, but continue to a greater value, known as the final size proportion, that has no close form solution, but can implicitly be defined as

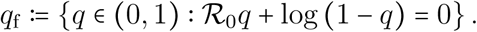

Finally, current prevalence in the community is simply given by

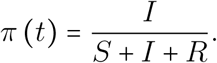

#### S2.9 Continuous time model

The Markovian continuous-time model analogous to S-1 corresponds to the following set of ordinary differential equations

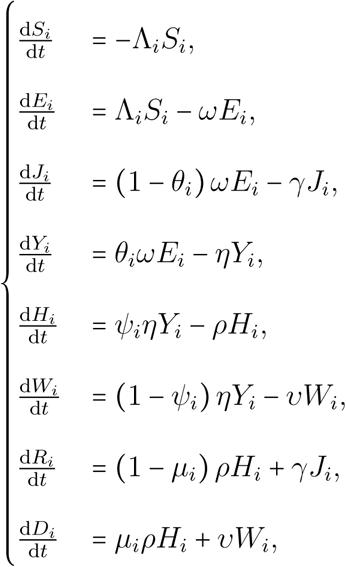

where the force of infection is given by

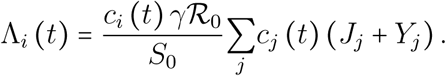

Note that a latent compartment *E* has been added to account for a delay between contamination time and the beginning of the infectious period. The average latency and infectious period are equal to *ω*^−1^ and *γ*^−1^. By construction, all transition times are exponentially distributed.

To compare this model to the focal non-Markovian discrete-time model introduced in this work, the output of the numerical integration was sampled at integer-valued time points. Then the fitting procedure used for the focal model was applied, though with a supplementary degree of freedom. Indeed, in the one hand, H is here exponentially distributed, therefore 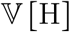 is determined by 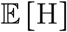 and, in the other hand, proper parameters *ω* and *γ* need to be fitted as there is no one-way relationship from generation time to latency and contagious periods. For information purposes, the maximum likelihood estimates and corresponding 95%-likelihood intervals of 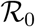, *t*_0_ and *κ* found by the estimation procedure applied to this Markovian model were respectively 4.3 [2.9, 5.8], 01 − 22 [01 − 20,01 − 23] and 26% [5,47] %.

### S3 Supplementary Results

#### S3.1 Maximum likelihood parameter estimates

**Table S-5:** Maximum likelihood estimates and associated 95% - likelihood intervals for the ten input parameters. Details about the estimation procedure are provided in section S2.7.

| main input parameter | notation | maximum likelihood estimates | 95% - likelihood interval |
| --- | --- | --- | --- |
| basic reproduction number | $\mathcal{R}_0$ | 2.99 | [2.59, 3.39] |
| initiation day (YY-MM-DD) | $t_0$ | 20-01-20 | [20-01-12, 20-01-28] |
| lock-down control (%) | $\kappa$ | 75.9 | [72.9, 78.7] |
| critical case contamination to hospitalization interval expectation (days) | $\mathbb{E}[H]$ | 14.5 | [13.6, 15.4] |
| critical case contamination to hospitalization interval variance (days<sup>2</sup>) | $\mathbb{V}[H]$ | 20.0 | [11.4, 30.9] |
| long ICU stay length expectation (days) | $\mathbb{E}[P]$ | 16.7 | [14.9, 18.8] |
| critical case hospitalization to death interval expectation (non long-stay ICU patients (days) | $\mathbb{E}[\Upsilon]$ | 6.63 | [6.19, 7.10] |
| infection fatality rate correction factor (%) | $\mathfrak{C}_F$ | 87.2 | [85.8, 88.5] |
| long-stay ICU fatality rate correction factor | $\mathfrak{C}_M$ | 100.3 | [100.2, 100.5] |
| long-stay ICU frequency correction factor | $\mathfrak{C}_\Psi$ | 93.8 | [93.0, 94.5] |

#### S3.2 Data right censoring and parameter inference

**Figure S-3:**
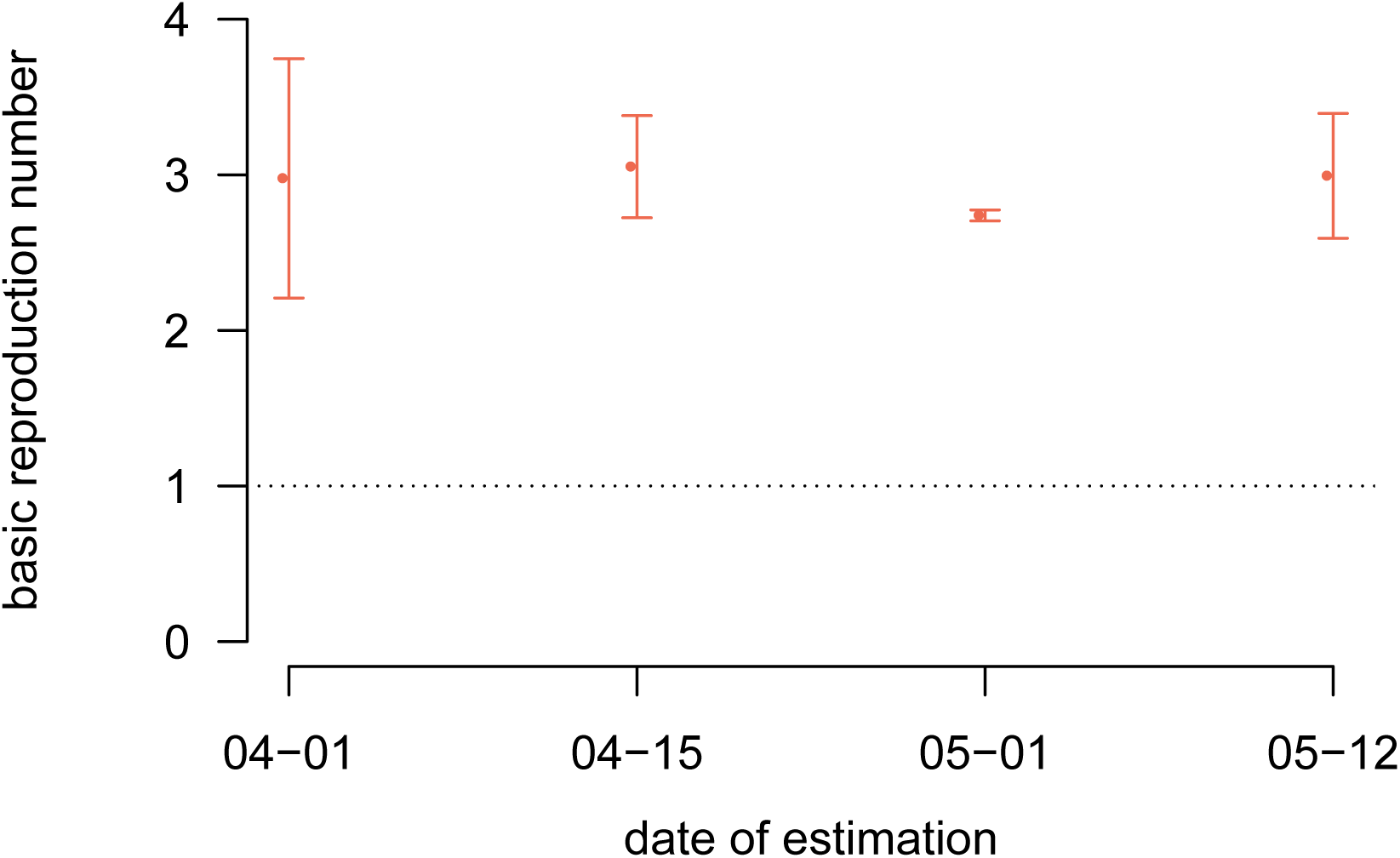
Basic reproduction number estimates as a function of the date of censoring. Each dot represents the maximum likelihood estimate of the basic reproduction number calculated from data truncated at the corresponding date on the x-axis. Bars shows 95% likelihood intervals.

**Figure S-4:**
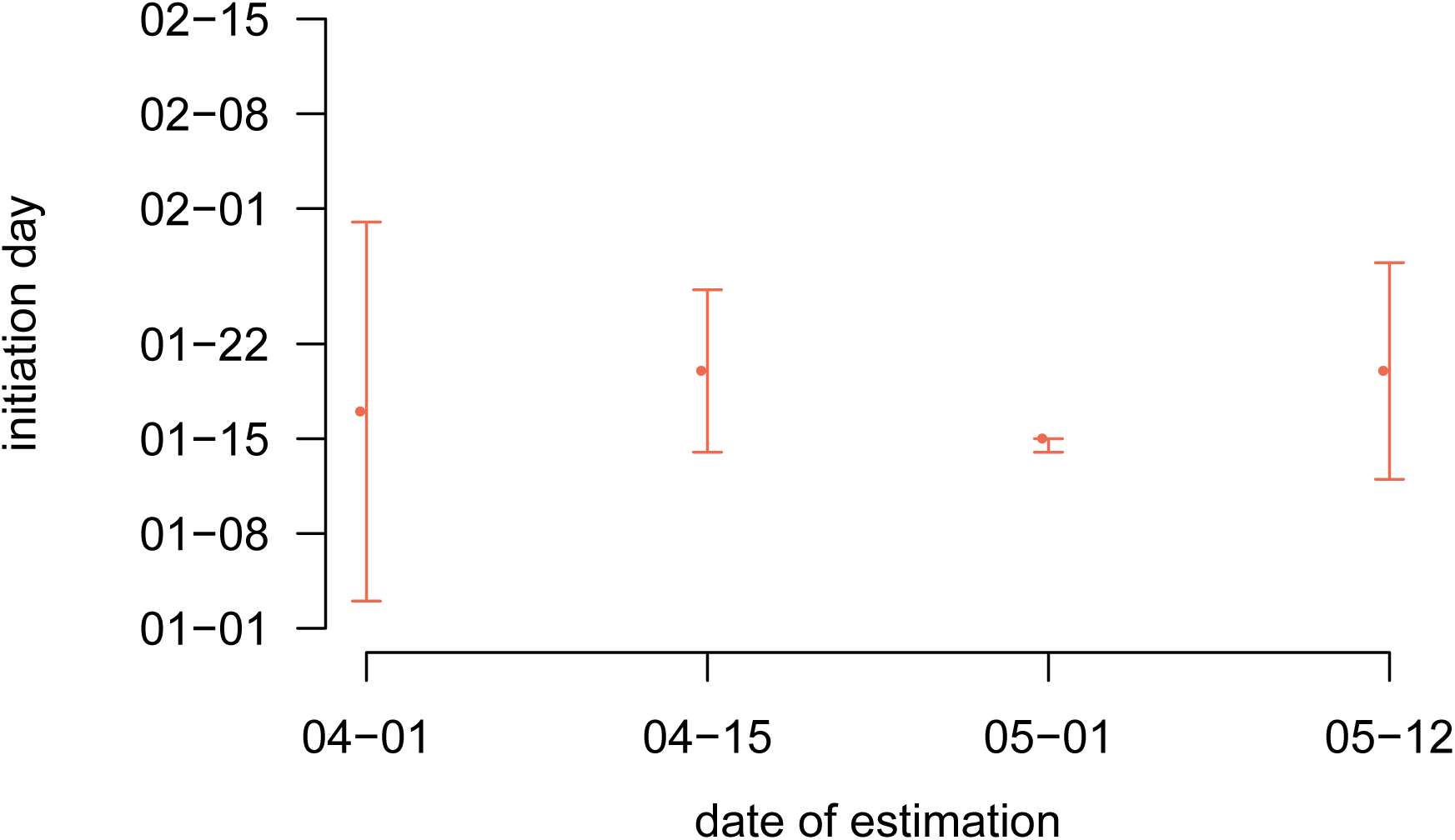
Successive initiation date estimates. Each dot represents the maximum likelihood estimate of the initiation date of the epidemic from data truncated at the corresponding date on the x-axis. Bars shows 95% likelihood intervals.

**Figure S-5:**
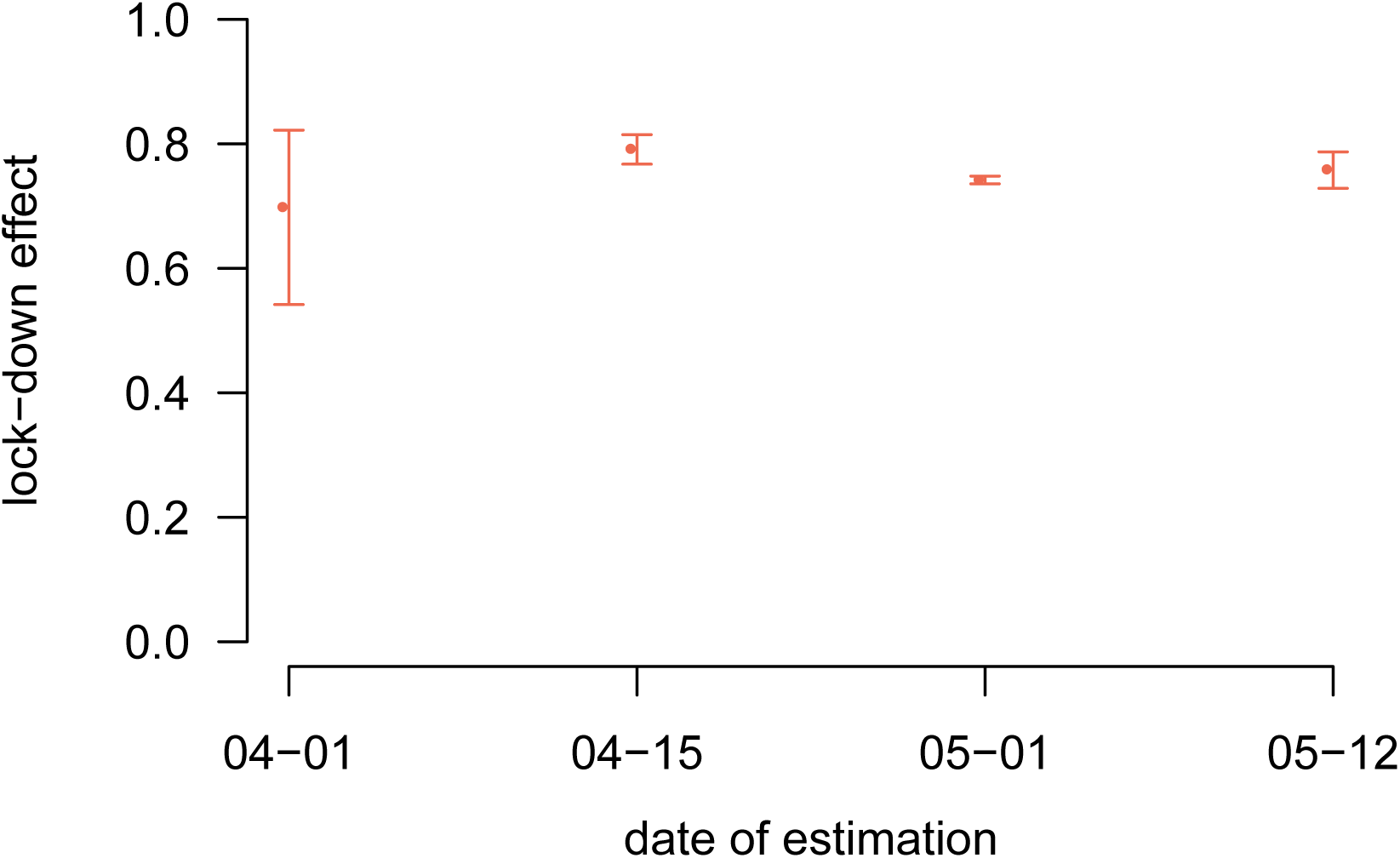
Successive lock-down effect estimates. Each dot represents the maximum likelihood estimate of the lock-down effect from data truncated at the corresponding date on the x-axis. Bars shows 95% likelihood intervals.

#### S3.3 Lock-down implementation date and cumulative mortality

**Figure S-6:**
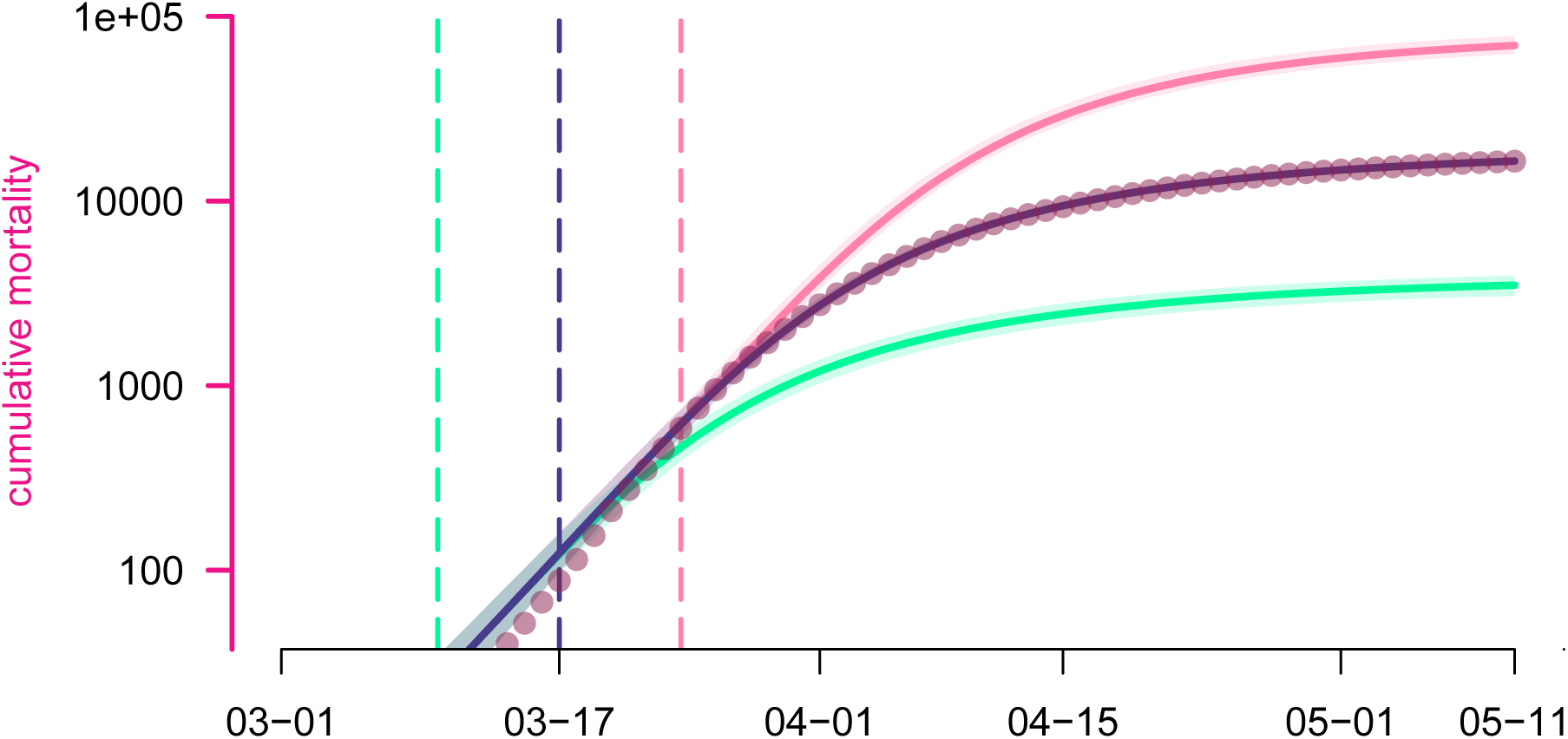
Estimated lock-down date effect on cumulative mortality. Each curve represents the median cumulative (hospital) mortality as generated by the model according to a given lock-down scenario, while their surrounding shaded areas correspond the their 95% confidence intervals. The scenarios are as detailed in Fig 4. Dots represent the data.

#### S3.4 Examples of post-May 11 scenarios

**Figure S-7:**
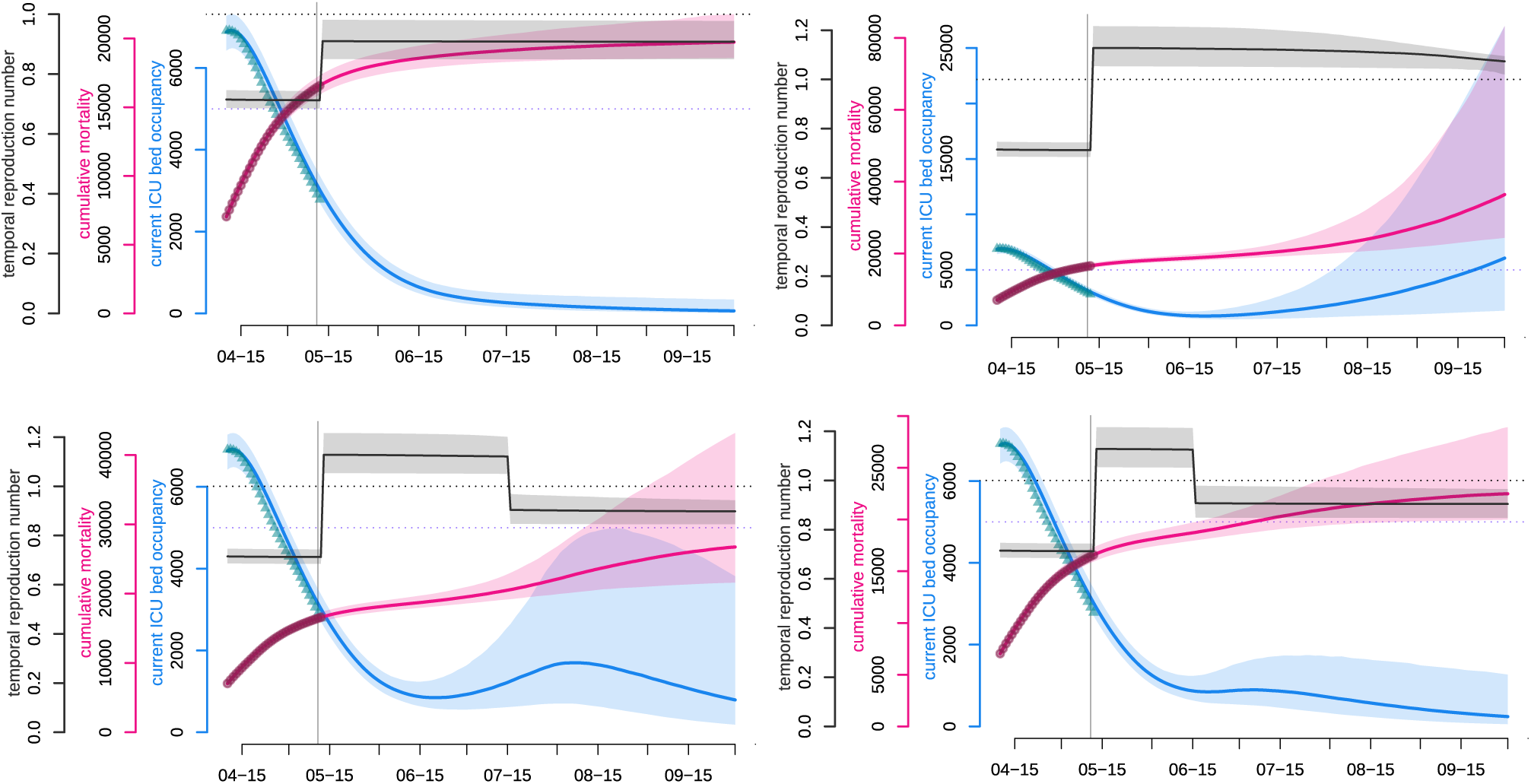
Examples of post-May 11 scenarios. Here are shown four runs of the model with distinct reproduction number after lock-down lifting and control response timings. Importantly, these plots are not statistical predictions of the future but only illustration-purpose qualitative explorations. The blue and pink curves respectively represent the median number of occupied beds in ICU nationwide and the median cumulative (hospital) mortality as generated by the fitted model. The turquoise triangles and red circles are the (rolling 7-day average) data counterparts. The black curve shows the median daily temporal reproduction number calculated from the simulated epidemic. The dotted horizontal line shows the reproduction number threshold value, i.e. 1. The purple dotted horizontal line shows the initial French ICU capacity, ca. 5,000 beds. The vertical corresponds to the end of the French national lock-down (May 11). Shaded areas correspond to 95% confidence intervals Note: the abscissa scales differ across panels. **Top left panel**. After lock-down lifting, the reproduction number 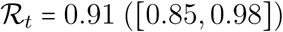, the epidemic is under control and vanishes spontaneously. **Top right panel**. After lock-down lifting, the reproduction number 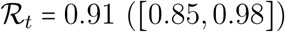, a second wave arises in absence of any control response. **Bottom left panel**. As previously, but a reinforced set of NPIs is implemented by Jul 15, reducing the reproduction number below 1. **Bottom right panel**. As previously, but a reinforced set of NPIs is implemented earlier, by Jun 15.

#### S3.5 Time vs intensity trade-off of the relaxed phase in the periodic lock-down strategy

**Figure S-8:**
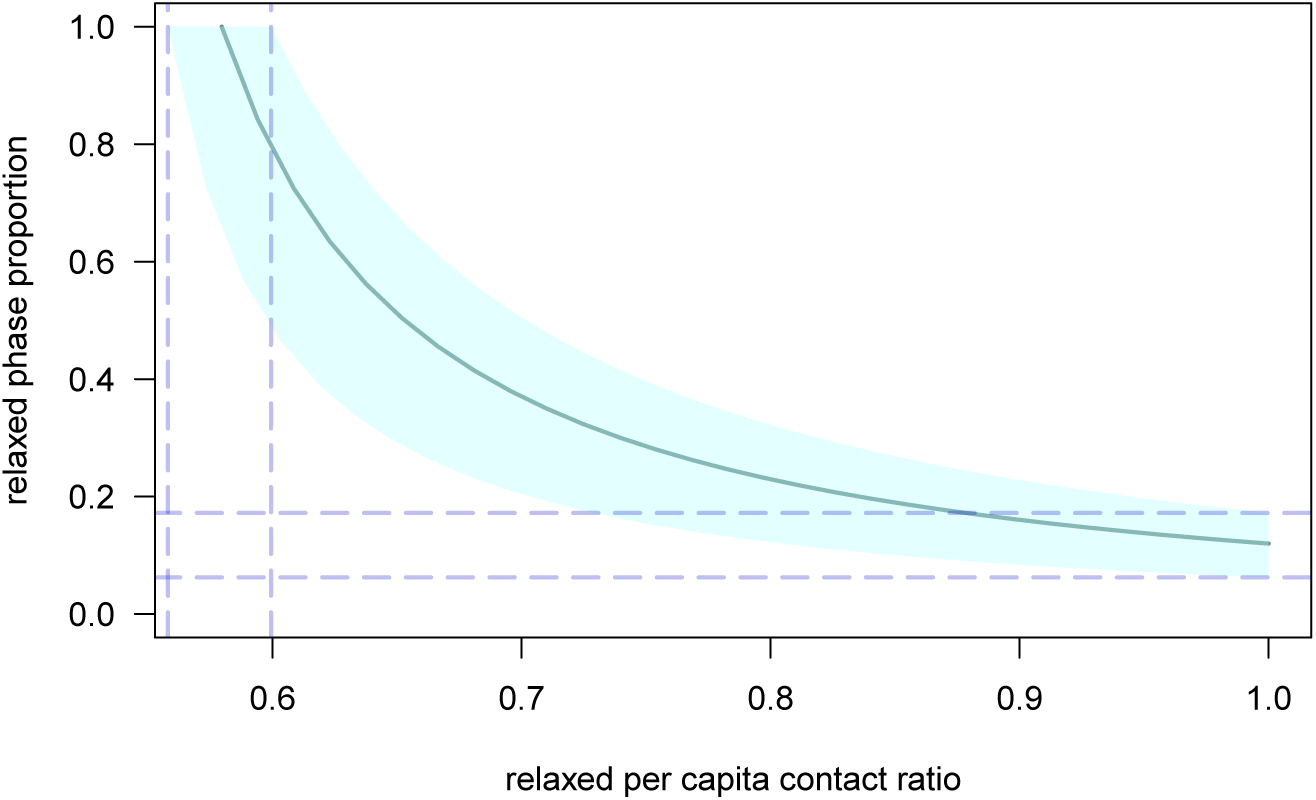
Time spent in the relaxed phase a function of the corresponding per capita contact ratio, in a periodic lock-down strategy. The maximum proportion of time possible to be spent in the relaxed phase of a NPI cycle without triggering a new epidemic, denoted by *p*_r,max_, is governed by three quantities, following eq.3: the basic reproduction number, 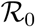, the per capita contact ratio during the hard phase of the NPI cycle (here set to the same value as during lock-down, *κ*), and the per capita contact ratio during the relaxed phase, *c*_r_. This figure illustrates the trade-off relation between *p*_r,max_ and *c*_r_: the central curve represents the median relation while the surrounding shaded areas correspond to the 95% confidence interval. Vertical dashed bars indicate the 95% CI of the average per capita contact ratio threshold associated to 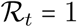 (therefore applicable the whole time, without need of any harder phase), this interval is [56,60] %. Horizontal dashed bars indicate the 95% of the proportion of the relaxed phase in each NPI cycle that guarantees associated to 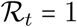, this interval is [6.2,12]%.

